# Mental health, gender, and care-seeking behavior during the COVID-19 pandemic in Sweden: An exploratory study

**DOI:** 10.1101/2023.02.08.23285645

**Authors:** Katalin Vincze, Gillian Murphy, Mary Barker, Juan González-Hijón, Anna K. Kähler, Emma M. Frans, Patrick F. Sullivan, Unnur A. Valdimarsdóttir, Fang Fang, Anikó Lovik

## Abstract

**Objective:** To explore the prevalence of care-seeking avoidance behavior in relation to gender and to describe the effect of (and potential interaction between) gender and care-seeking on mental health during the COVID-19 pandemic in Sweden.

**Methods:** We performed a cross-sectional study among 27,562 participants of the Omtanke2020 Study, using data collected at three time points concerning sociodemographic factors, mental health symptoms, and care-seeking behavior. Network analysis and prevalence ratios calculated from modified Poisson regressions were used to explore the relationship between gender, care-seeking behavior, and mental health symptoms (depression, anxiety, and COVID-19-related distress).

**Results:** In our study, women reported a higher prevalence of mental health symptoms and avoidance of care-seeking due to COVID-19, compared to men. At baseline and six months thereafter, female gender was positively associated with COVID-19-related distress and previous mental health diagnosis. At 12 months after baseline, female gender was positively associated with anxiety and avoidance of care-seeking for mental health. However, previous mental health diagnosis and care avoidance were more strongly associated with a higher prevalence of mental health symptoms among men, compared to women.

**Conclusion:** This study highlights gender differences in mental health outcomes and care-seeking behavior during the COVID-19 pandemic in Sweden.

**Funding:** This work was supported with grants from Nordforsk (COVIDMENT, 105668 and 138929).

## Introduction

Gender has consistently presented as an important determinant of both physical and mental health (1–3). There are gender differences in risk factors, prevalence, symptomatology, and prognosis of mental health (2,3). The extent of healthcare utilization is another important factor, which can impact on individual ‘s mental health trajectory. While many studies have found that women typically seek care for both physical and mental health concerns more often than men (4–7), much of this evidence comes from cross-sectional studies, with results varying depending on the setting and outcomes being assessed (8–11).

The impact of the COVID-19 pandemic has been far-reaching, with consequences on mental health and care-seeking behavior. During the early stages of the pandemic (i.e., February-September 2020), several cross-sectional studies from various settings showed that many adults avoided seeking medical care, either urgent or routine, as well as care for pre-existing or newly-onset psychological symptoms (12–16). Data from the United States suggests that delaying care multiple times during the pandemic led to more adverse outcomes than delaying care once or not at all (17). Sweden has a decentralized, publicly funded healthcare system with universal coverage for all residents (18,19) and existing e-Health mental health solutions (20,21). Nevertheless, insufficient access to care has been reported among socioeconomically disadvantaged groups, which has only been exacerbated by the pandemic (22,23). Sweden had very different COVID-19 prevention strategy in the first wave of the pandemic from other Nordic and European countries, with a range of recommendations for social distancing rather than strict lockdown measures (24). Regardless, there is evidence that Swedish residents still suffered from negative physical and mental health outcomes during the pandemic (20,25,26).

Data on the COVID-19 pandemic and the impact it has had on care-seeking behavior tends to come from cross-sectional studies (12–16). There is currently a shortage of studies with longitudinal data and studies focusing on potential gender difference. Therefore, we aimed to first describe how care-seeking behavior differed between men and women over three time points during the pandemic in Sweden. Secondly, we aimed to describe the relationships between mental health, gender, care-seeking, and delayed care at all three time points. Finally, we aimed to investigate the association between care-seeking and mental health, by gender.

## Methods

### Study design

This study used data from participants of the Omtanke2020 Study, an ongoing longitudinal cohort study initiated in June 2020 (26). Details of the study design of Omtanke2020 have previously been published (26). The Omtanke2020 Study was approved by the Swedish Ethics Authority (no. 2020– 01785) and all participants provided written informed consent (27). In the present study, we used data collected at three time points, namely baseline, 6-month follow-up, and 12-month follow-up. Baseline data collection was carried out between 9 June 2020 - 8 June 2021. The 6-month follow-up occurred approximately six months after the participants completed the baseline questionnaire (mostly March-November 2021) (26). The 12-month follow-up was carried out 12-18 months after the participants completed the baseline questionnaire (mostly December 2021 – February 2022).

### Variables

Age at baseline and gender (male/female) were collected through the unique Swedish personal identity numbers (PIN) the participants used to access the study questionnaires (26,28). Sweden currently only recognizes male and female as genders (29).

Somatic health (SH) and mental health (MH) were distinguished in the questionnaires, with somatic health referred to as physical health. General self-reported somatic and mental health, as well as disease history, were measured with separate questions (“How do you rate your physical/mental health in general? Very good, good, decent, bad”). Participants always had the response option “I cannot/do not want to answer” when filling out the questionnaires, resulting in missing values.

Care-seeking was identified by asking participants whether they avoided care-seeking for mental health due to worries of getting COVID-19 (yes, no), whether they avoided care-seeking for somatic health due to worries of getting COVID-19 (yes, no), whether they had delayed care (yes, no), how long the delay was (less than a month, one to four months, more than four months, unspecified delay, cancelled), the type of care delayed (cancer treatment, operation, X-ray examination including magnetic resonance and computerized tomography, visit at the general practice, other care) and the level of worry caused by the delay in care (very much, quite a lot, neutral, quite little, not worried at all).

Mental health symptoms (depression, anxiety, and COVID-19 related distress) were measured using validated questionnaires.

Depression was measured using the Patient Health Questionnaire (PHQ-9), consisting of nine items (30). Internal consistency for PHQ-9 measurement in the baseline sample of Omtanke2020 was reported to be α = 0.88 (26). Scores for the PHQ-9 can be categorized as minimal (scores of 0-4), mild (5-9), moderate (10-14), moderately severe (15-20) and severe (scores of 21-27) (26,30). When treated as a binary variable, the recommended score of ≥10 was used as cut-off (30,31).

The Generalized Anxiety Disorder 7-item scale (GAD-7) was used to measure anxiety (32). Cronbach ‘s alpha for the GAD-7 measurement in the baseline sample of Omtanke20202 was reported to be α = 0.90 (26). Categorization of scores include minimal (scores of 0-4), mild (5-9), moderate (10-14), and severe (15-21) (26,32). When used as a binary variable, the recommended score of ≥10 was used as cut-off (31,32).

COVID-19-related distress symptoms were measured using a modified, five-item version of the Primary Care PTSD Screen for DSM-5 (PC-PTSD-5) scale (33). Internal consistency for this measure in the baseline Omtanke2020 sample was α = 0.77 (26). The cut-off of ≥4, as suggested in previous literature (31,33) was used to define a binary variable.

### Statistical analysis

All sociodemographic, mental health and care-seeking variables were first summarized using mean (standard deviation) for continuous variables or frequencies (percentages) for categorical variables.

### Mixed Graphical Model (MGM)

A network analysis was performed to visualize the relationships between the following variables at baseline, 6-month follow-up, and 12-month follow-up: gender, age, mental health symptoms (depression, anxiety, COVID-19-related distress), avoidance of care-seeking for somatic health, avoidance of care-seeking for mental health, delayed care, presence of somatic comorbidities, previous mental health diagnosis, and cumulative COVID-19 status. Mixed graphical models (MGMs) were plotted using the *mgm package in R* (34). MGMs allow for appropriate correlations to be run between variables of different types (count, categorical, continuous) (34–36).

### Prevalence ratios of depression, anxiety, and COVID-19-related distress

To investigate the association between care-seeking behavior and mental health, prevalence ratios were calculated using modified robust Poisson regression at the three time points (37–39), with three different models performed at each time point. The default model included age, previous mental health diagnosis, cumulative COVID-19 status, experience of delayed care, avoidance of care-seeking for mental health, and avoidance of care-seeking for somatic health. Model 1 additionally included an interaction between avoidance of care-seeking for mental health and gender. Model 2 included an interaction between avoidance of care-seeking for somatic health and gender. Model 3 included n interaction between delayed care and gender. Stratification by gender was performed for models with a significant interaction effect. The *glm* function from the R *stats* package was used to perform the models (40).

All analyses were conducted using R (version 4.2.1) (40).

## Results

Table 1 summarizes sociodemographic characteristics and mental and physical health of the participants at baseline. Majority of the participants (81.47%) were female. The mean age was 48.76 years (SD = 15.73). Most participants (72.44%) reported being in a relationship. Approximately half of the participants (50.69%) reported a BMI in the normal range, whilst 51.84% had never smoked. Over half of the participants did not engage in habitual drinking. The majority of participants reported no previous mental health diagnosis (66.44%). However, a high frequency of mental health symptoms was reported by participants, with higher percentages observed among women (12.59% vs 10.03% for moderate depressive symptoms, 6.21% vs 4.76% for moderately severe depressive symptoms, 3.10% vs 2.72% for severe depressive symptoms, 9.58% vs 7.23% for moderate anxiety symptoms, 6.04% vs 4.05% for severe anxiety symptoms, and 32.40% vs 19.05% for severe COVID-19-related distress). The majority of participants reported no somatic disease at baseline (66.25%). At baseline, 5.18% of participants reported that they had been diagnosed with COVID-19, which accumulated to 42.99% at 12-month follow-up.

**Table 1:** Baseline characteristics of the participants

| Baseline characteristics | Total<br>(n=27562) | Male<br>(n=5107) | Female<br>(n=22455) |
| --- | --- | --- | --- |
| **Age, years** |  |  |  |
| Mean (SD) | 48.76 (15.73) | 47.53 (16.46) | 49.04 (15.55) |
| **Age group, N (%)** |  |  |  |
| 18-29 | 3769 (13.67) | 793 (15.53) | 2976 (13.25) |
| 30-39 | 5016 (18.20) | 1097 (21.48) | 3919 (17.45) |
| 40-49 | 5274 (19.14) | 1024 (20.05) | 4250 (18.93) |
| 50-59 | 5932 (21.52) | 843 (16.51) | 5089 (22.66) |
| 60-69 | 4404 (15.98) | 695 (13.61) | 3709 (16.52) |
| 70+ | 3167 (11.49) | 655 (12.83) | 2512 (11.9) |
| **Relationship status, N (%)** |  |  |  |
| In a relationship | 19966 (72.44) | 3793 (74.27) | 16173 (72.02) |
| Single | 7459 (27.06) | 1292 (25.30) | 6167 (27.46) |
| Missing | 137 (0.50) | 22 (0.43) | 115 (0.51) |
| **Body Mass Index, N (%)** |  |  |  |
| Underweight (<18.5) | 1660 (6.02) | 147 (2.88) | 1513 (6.74) |
| Normal weight (18.5-25) | 13972 (50.69) | 2457 (48.11) | 11515 (51.28) |
| Overweight (25-30) | 8162 (29.61) | 1871 (36.64) | 6291 (28.02) |
| Obese (>30) | 3768 (13.67) | 632 (12.38) | 3136 (13.97) |
| Missing | 1208 (4.38) | 82 (1.61) | 1126 (5.01) |
| **Smoking or snuff, N (%)** |  |  |  |
| Never | 14287 (51.84) | 2442 (47.82) | 11845 (52.75) |
| Formerly | 4645 (16.85) | 1295 (25.36) | 3350 (14.92) |
| Current | 8456 (30.68) | 1341 (26.26) | 7115 (31.69) |
| Missing | 174 (0.63) | 29 (0.57) | 145 (0.65) |
| **Binge drinking, N (%)** |  |  |  |
| No | 14943 (54.22) | 2838 (55.57) | 12105 (53.91) |
| Yes | 7268 (26.37) | 1385 (27.12) | 5883 (26.20) |
| Missing | 5351 (19.41) | 884 (17.31) | 4467 (19.89) |
| **Previous mental health diagnosis, N (%)** |  |  |  |
| No | 17760 (64.44) | 3831 (75.01) | 13929 (62.03) |
| Yes | 9438 (34.24) | 1218 (23.85) | 8220 (36.61) |
| Missing | 364 (1.32) | 58 (1.14) | 306 (1.36) |
| **Mental and physical health characteristics** |  |  |  |
| **Mental health scales, mean (SD)** |  |  |  |
| PHQ-9 | 5.793 (5.49) | 5.156 (5.34) | 5.938 (5.51) |
| GAD-7 | 4.405 (4.77) | 3.67 (4.40) | 4.572 (4.84) |
| Modified PC-PTSD-5 | 2.388 (1.59) | 1.885 (1.52) | 2.503 (1.58) |
| General mental health | 2.147 (0.916) | 2.09 (0.94) | 2.16 (0.91) |
| General physical health | 2.094 (0.84) | 2.050 (0.85) | 2.100 (0.84) |
| **Symptoms of depression, N (%)** |  |  |  |
| Minimal (0-4) | 14451 (52.43) | 2938 (57.53) | 11513 (51.27) |
| Mild (5-9) | 7298 (26.48) | 1275 (24.97) | 6023 (26.82) |
| Moderate (10-14) | 3339 (12.11) | 512 (10.03) | 2827 (12.59) |
| Moderately severe (15-20) | 1638 (5.94) | 243 (4.76) | 1395 (6.21) |
| Severe (21-27) | 836 (3.03) | 139 (2.72) | 697 (3.10) |
| **Symptoms of anxiety, N (%)** |  |  |  |
| Minimal (0-4) | 17339 (62.91) | 3565 (69.81) | 13774 (61.34) |
| Mild (5-9) | 6139 (22.27) | 966 (18.92) | 5173 (23.04) |
| Moderate (10-14) | 2521 (9.15) | 369 (7.23) | 2152 (9.58) |
| Severe (15-21) | 1563 (5.67) | 207 (4.05) | 1356 (6.04) |
| **Symptoms of COVID-19-related distress, N (%)** |  |  |  |
| None (0) | 4387 (15.92) | 1181 (23.13) | 3206 (14.28) |
| Moderate (1-3) | 14371 (52.14) | 2879 (56.37) | 11492 (51.18) |
| Severe (4-5) | 8248 (29.93) | 973 (19.05) | 7275 (32.40) |
| Missing | 556 (2.02) | 74 (1.45) | 482 (2.15) |
| **General mental health, N (%)** |  |  |  |
| Very good | 7531 (27.32) | 1602 (31.37) | 5929 (26.40) |
| Quite good | 10728 (38.92) | 1895 (37.11) | 8833 (39.34) |
| Decent | 6996 (25.38) | 1158 (22.67) | 5838 (26.00) |
| Bad | 2295 (8.33) | 452 (8.85) | 1843 (8.21) |
| Missing | 12 (0.04) | 0 | 12 (0.05) |
| **Number of somatic diseases, N (%)** |  |  |  |
| 0 | 18259 (66.25) | 3334 (65.28) | 14925 (66.47) |
| 1 | 6473 (23.49) | 1154 (22.60) | 5319 (23.69) |
| 2 | 1724 (6.25) | 380 (7.44) | 1344 (5.99) |
| ≥3 | 480 (1.74) | 104 (2.04) | 376 (1.67) |
| Missing | 626 (2.27) | 135 (2.64) | 491 (2.19) |
| **General physical health, N (%)** |  |  |  |
| Very good | 7079 (25.68) | 1457 (28.53) | 5622 (25.04) |
| Quite good | 12234 (44.39) | 2186 (42.80) | 10048 (44.75) |
| Decent | 6825 (24.76) | 1215 (23.79) | 5610 (24.98) |
| Bad | 1416 (5.14) | 247 (4.84) | 1169 (5.21) |
| Missing | 8 (0.03) | 2 (0.04) | 6 (0.03) |
| **Cumulative positive COVID-19 status, N (%)** |  |  |  |
| Baseline | 1428 (5.18) | 305 (5.97) | 1123 (5) |
| **Self-recruitment, N (%)** |  |  |  |
| No | 12052 (43.73) | 2373 (46.47) | 9679 (43.10) |
| Yes | 11353 (41.19) | 2336 (45.74) | 9017 (40.16) |
| Missing | 4157 (15.08) | 398 (7.79) | 3759 (16.74) |

Note: SD = Standard deviation, PHQ-9 = Patient Health Questionnaire (measuring symptoms of depression), GAD-7 = Generalized Anxiety Disorder 7-item scale, PC-PTSD-5 = Primary Care PTSD Screen for DSM-5 (measuring COVID-19-related distress through modification)

A higher percentage of women, compared to men, avoided seeking care due to COVID-19, both for mental (3.99% vs 3.41% at baseline, 1.40% vs.0.70% at 6-month follow-up, 1.26% vs 1.06% at 12-month follow-up) and somatic health (14.31% vs 10.51% at baseline, 5.53% vs 2.72% at 6-month follow-up, and 4.47% vs 3.09% at 12-month follow-up) (Table2). Women also reported more delayed care (16.45% vs 12.69% at baseline, 5.50% vs 3.64% at 6-month follow-up, 8.27% vs 6.07% at 12-month follow-up). However, at the two follow-up time points, men more frequently had missing responses. A small proportion of participants reported worries about delayed care, with under 1% of participants reporting that they were ‘very much ‘ worried.

### Mixed Graphical Models (MGM)

The networks shown in Figure 1 illustrate that anxiety and depression were highly positively correlated at all three time points. At both baseline and 6-month follow-up, female gender was positively associated with COVID-19-related distress and previous mental health diagnosis. At 12-month follow-up, female gender was additionally associated with anxiety and avoidance of care-seeking for mental health. Avoidance of care-seeking for mental and somatic health due to worry of COVID-19 was strongly positively correlated at all time points. A negative correlation between delayed care and avoidance of care-seeking for somatic health was also found at all three time points.

**Figure 1:**
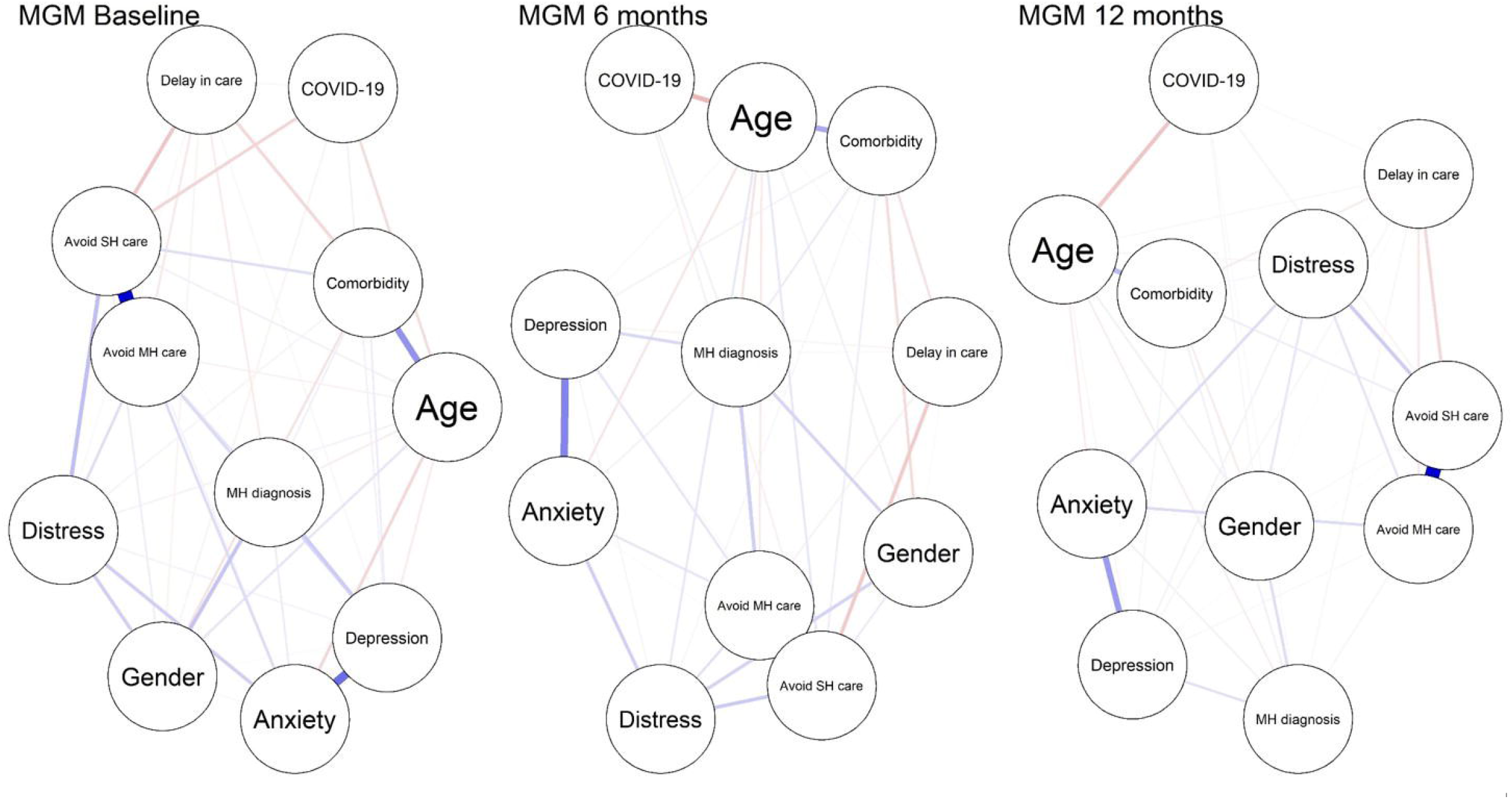
Mixed graphical models (MGMs) Note: ‘Avoid MH care ‘ stands for having previously avoided care-seeking for mental health due to reasons related to the COVID-19 pandemic, ‘Avoid SH care ‘ stands for the same but with somatic health, ‘Comorbidity ‘ stands for somatic comorbidities, ‘COVID-19 ‘ stands for cumulative COVID-19 diagnosis, ‘Distress ‘ stands for COVID-19-related distress and ‘MH diagnosis ‘ stands for previous mental health diagnosis.

### Prevalence ratios

Female gender was associated with higher prevalence of depression, anxiety, and COVID-19-related distress (Table 3). Age and cumulative COVID-19 status were associated with a lower prevalence, whereas previous mental health diagnosis, experience of delayed care, avoidance of care-seeking for mental health, and avoidance of care-seeking for somatic health were associated with a higher prevalence. In the gender-stratified analyses, we found however that a previous mental health diagnosis was more strongly associated with the risk of depression, anxiety, or COVID-19-related distress at the time of data collection, among men compared to women. Similarly, the associations of avoidance of care-seeking for mental health and somatic health, as well as experiencing delayed care, with the risk of depression, anxiety, and COVID-19-related distress were also stronger among men compared to women.

**Table 2:**
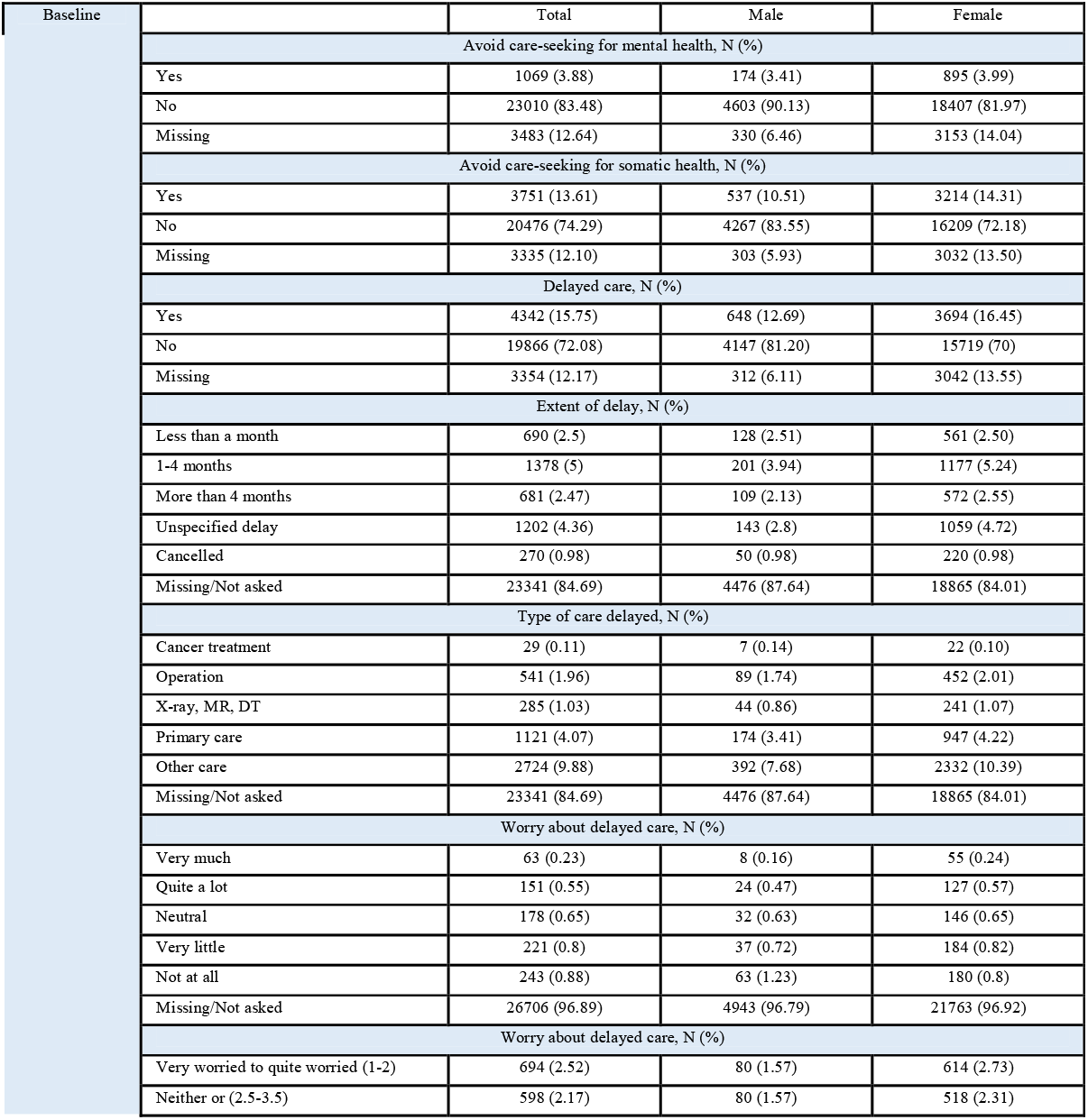
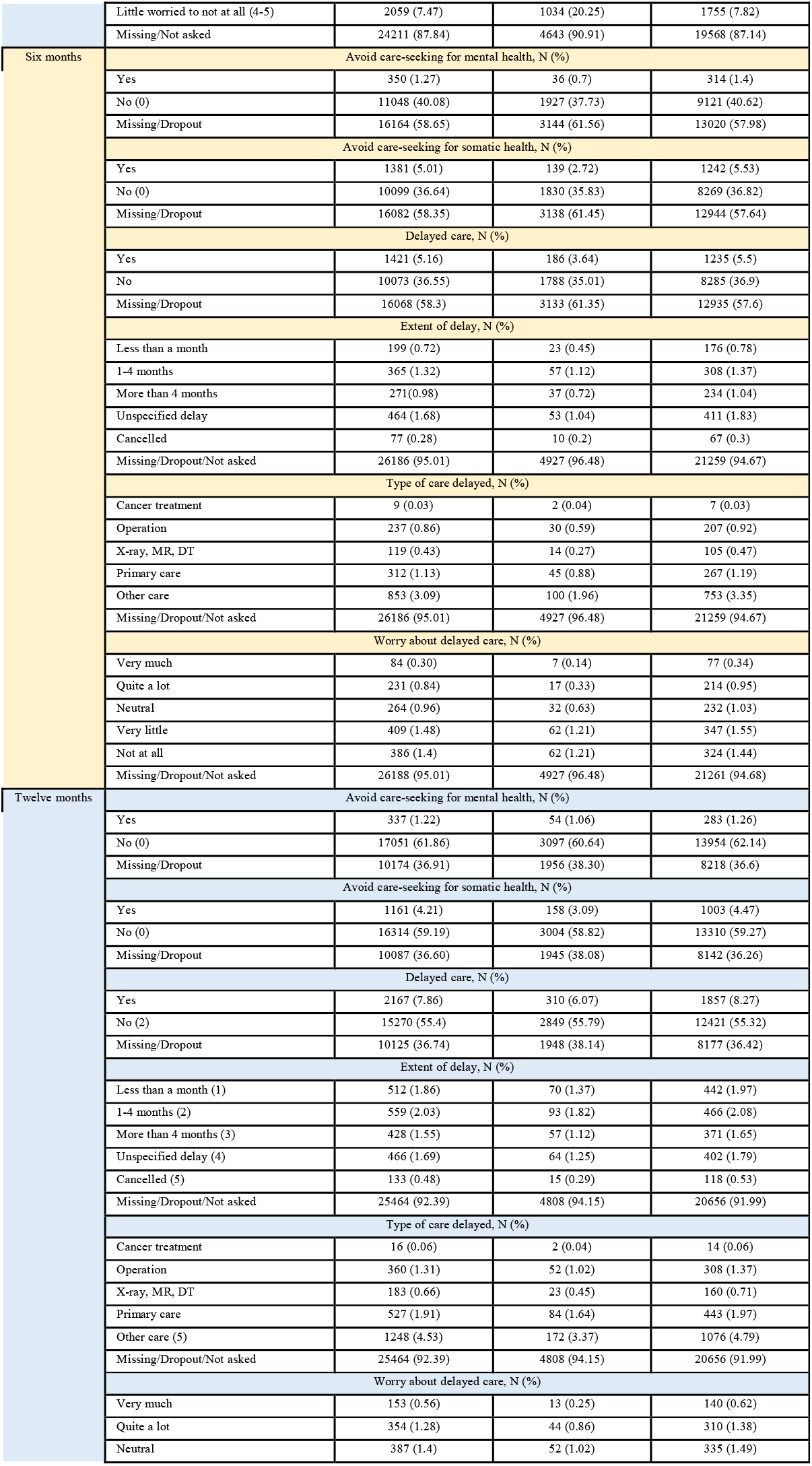
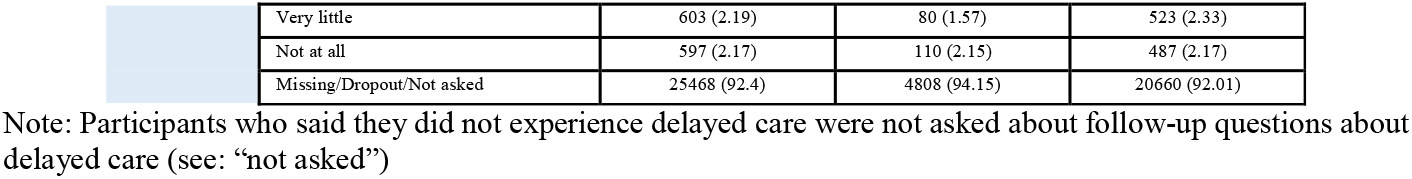
Care-seeking and delayed care at 3 timepoints, by gender

| Baseline | Total | Male | Female |
| --- | --- | --- | --- |
| **Avoid care-seeking for mental health, N (%)** |  |  |  |
| Yes | 1069 (3.88) | 174 (3.41) | 895 (3.99) |
| No | 23010 (83.48) | 4603 (90.13) | 18407 (81.97) |
| Missing | 3483 (12.64) | 330 (6.46) | 3153 (14.04) |
| **Avoid care-seeking for somatic health, N (%)** |  |  |  |
| Yes | 3751 (13.61) | 537 (10.51) | 3214 (14.31) |
| No | 20476 (74.29) | 4267 (83.55) | 16209 (72.18) |
| Missing | 3335 (12.10) | 303 (5.93) | 3032 (13.50) |
| **Delayed care, N (%)** |  |  |  |
| Yes | 4342 (15.75) | 648 (12.69) | 3694 (16.45) |
| No | 19866 (72.08) | 4147 (81.20) | 15719 (70) |
| Missing | 3354 (12.17) | 312 (6.11) | 3042 (13.55) |
| **Extent of delay, N (%)** |  |  |  |
| Less than a month | 690 (2.5) | 128 (2.51) | 561 (2.50) |
| 1-4 months | 1378 (5) | 201 (3.94) | 1177 (5.24) |
| More than 4 months | 681 (2.47) | 109 (2.13) | 572 (2.55) |
| Unspecified delay | 1202 (4.36) | 143 (2.8) | 1059 (4.72) |
| Cancelled | 270 (0.98) | 50 (0.98) | 220 (0.98) |
| Missing/Not asked | 23341 (84.69) | 4476 (87.64) | 18865 (84.01) |
| **Type of care delayed, N (%)** |  |  |  |
| Cancer treatment | 29 (0.11) | 7 (0.14) | 22 (0.10) |
| Operation | 541 (1.96) | 89 (1.74) | 452 (2.01) |
| X-ray, MR, DT | 285 (1.03) | 44 (0.86) | 241 (1.07) |
| Primary care | 1121 (4.07) | 174 (3.41) | 947 (4.22) |
| Other care | 2724 (9.88) | 392 (7.68) | 2332 (10.39) |
| Missing/Not asked | 23341 (84.69) | 4476 (87.64) | 18865 (84.01) |
| **Worry about delayed care, N (%)** |  |  |  |
| Very much | 63 (0.23) | 8 (0.16) | 55 (0.24) |
| Quite a lot | 151 (0.55) | 24 (0.47) | 127 (0.57) |
| Neutral | 178 (0.65) | 32 (0.63) | 146 (0.65) |
| Very little | 221 (0.8) | 37 (0.72) | 184 (0.82) |
| Not at all | 243 (0.88) | 63 (1.23) | 180 (0.8) |
| Missing/Not asked | 26706 (96.89) | 4943 (96.79) | 21763 (96.92) |
| **Worry about delayed care, N (%)** |  |  |  |
| Very worried to quite worried (1-2) | 694 (2.52) | 80 (1.57) | 614 (2.73) |
| Neither or (2.5-3.5) | 598 (2.17) | 80 (1.57) | 518 (2.31) |

|  |  |  |  |  |
| --- | --- | --- | --- | --- |
|  | Little worried to not at all (4-5) | 2059 (7.47) | 1034 (20.25) | 1755 (7.82) |
|  | Missing/Not asked | 24211 (87.84) | 4643 (90.91) | 19568 (87.14) |
| Six months | Avoid care-seeking for mental health, N (%) |  |  |  |
|  | Yes | 350 (1.27) | 36 (0.7) | 314 (1.4) |
|  | No (0) | 11048 (40.08) | 1927 (37.73) | 9121 (40.62) |
|  | Missing/Dropout | 16164 (58.65) | 3144 (61.56) | 13020 (57.98) |
|  | Avoid care-seeking for somatic health, N (%) |  |  |  |
|  | Yes | 1381 (5.01) | 139 (2.72) | 1242 (5.53) |
|  | No (0) | 10099 (36.64) | 1830 (35.83) | 8269 (36.82) |
|  | Missing/Dropout | 16082 (58.35) | 3138 (61.45) | 12944 (57.64) |
|  | Delayed care, N (%) |  |  |  |
|  | Yes | 1421 (5.16) | 186 (3.64) | 1235 (5.5) |
|  | No | 10073 (36.55) | 1788 (35.01) | 8285 (36.9) |
|  | Missing/Dropout | 16068 (58.3) | 3133 (61.35) | 12935 (57.6) |
|  | Extent of delay, N (%) |  |  |  |
|  | Less than a month | 199 (0.72) | 23 (0.45) | 176 (0.78) |
|  | 1-4 months | 365 (1.32) | 57 (1.12) | 308 (1.37) |
|  | More than 4 months | 271 (0.98) | 37 (0.72) | 234 (1.04) |
|  | Unspecified delay | 464 (1.68) | 53 (1.04) | 411 (1.83) |
|  | Cancelled | 77 (0.28) | 10 (0.2) | 67 (0.3) |
|  | Missing/Dropout/Not asked | 26186 (95.01) | 4927 (96.48) | 21259 (94.67) |
|  | Type of care delayed, N (%) |  |  |  |
|  | Cancer treatment | 9 (0.03) | 2 (0.04) | 7 (0.03) |
|  | Operation | 237 (0.86) | 30 (0.59) | 207 (0.92) |
|  | X-ray, MR, DT | 119 (0.43) | 14 (0.27) | 105 (0.47) |
|  | Primary care | 312 (1.13) | 45 (0.88) | 267 (1.19) |
|  | Other care | 853 (3.09) | 100 (1.96) | 753 (3.35) |
|  | Missing/Dropout/Not asked | 26186 (95.01) | 4927 (96.48) | 21259 (94.67) |
|  | Worry about delayed care, N (%) |  |  |  |
|  | Very much | 84 (0.30) | 7 (0.14) | 77 (0.34) |
|  | Quite a lot | 231 (0.84) | 17 (0.33) | 214 (0.95) |
|  | Neutral | 264 (0.96) | 32 (0.63) | 232 (1.03) |
|  | Very little | 409 (1.48) | 62 (1.21) | 347 (1.55) |
|  | Not at all | 386 (1.4) | 62 (1.21) | 324 (1.44) |
|  | Missing/Dropout/Not asked | 26188 (95.01) | 4927 (96.48) | 21261 (94.68) |
| Twelve months | Avoid care-seeking for mental health, N (%) |  |  |  |
|  | Yes | 337 (1.22) | 54 (1.06) | 283 (1.26) |
|  | No (0) | 17051 (61.86) | 3097 (60.64) | 13954 (62.14) |
|  | Missing/Dropout | 10174 (36.91) | 1956 (38.30) | 8218 (36.6) |
|  | Avoid care-seeking for somatic health, N (%) |  |  |  |
|  | Yes | 1161 (4.21) | 158 (3.09) | 1003 (4.47) |
|  | No (0) | 16314 (59.19) | 3004 (58.82) | 13310 (59.27) |
|  | Missing/Dropout | 10087 (36.60) | 1945 (38.08) | 8142 (36.26) |
|  | Delayed care, N (%) |  |  |  |
|  | Yes | 2167 (7.86) | 310 (6.07) | 1857 (8.27) |
|  | No (2) | 15270 (55.4) | 2849 (55.79) | 12421 (55.32) |
|  | Missing/Dropout | 10125 (36.74) | 1948 (38.14) | 8177 (36.42) |
|  | Extent of delay, N (%) |  |  |  |
|  | Less than a month (1) | 512 (1.86) | 70 (1.37) | 442 (1.97) |
|  | 1-4 months (2) | 559 (2.03) | 93 (1.82) | 466 (2.08) |
|  | More than 4 months (3) | 428 (1.55) | 57 (1.12) | 371 (1.65) |
|  | Unspecified delay (4) | 466 (1.69) | 64 (1.25) | 402 (1.79) |
|  | Cancelled (5) | 133 (0.48) | 15 (0.29) | 118 (0.53) |
|  | Missing/Dropout/Not asked | 25464 (92.39) | 4808 (94.15) | 20656 (91.99) |
|  | Type of care delayed, N (%) |  |  |  |
|  | Cancer treatment | 16 (0.06) | 2 (0.04) | 14 (0.06) |
|  | Operation | 360 (1.31) | 52 (1.02) | 308 (1.37) |
|  | X-ray, MR, DT | 183 (0.66) | 23 (0.45) | 160 (0.71) |
|  | Primary care | 527 (1.91) | 84 (1.64) | 443 (1.97) |
|  | Other care (5) | 1248 (4.53) | 172 (3.37) | 1076 (4.79) |
|  | Missing/Dropout/Not asked | 25464 (92.39) | 4808 (94.15) | 20656 (91.99) |
|  | Worry about delayed care, N (%) |  |  |  |
|  | Very much | 153 (0.56) | 13 (0.25) | 140 (0.62) |
|  | Quite a lot | 354 (1.28) | 44 (0.86) | 310 (1.38) |
|  | Neutral | 387 (1.4) | 52 (1.02) | 335 (1.49) |
| Very little | 603 (2.19) | 80 (1.57) | 523 (2.33) |
| Not at all | 597 (2.17) | 110 (2.15) | 487 (2.17) |
| Missing/Dropout/Not asked | 25468 (92.4) | 4808 (94.15) | 20660 (92.01) |

Note: Participants who said they did not experience delayed care were not asked about follow-up questions about delayed care (see: “not asked”)

**Table 3:**
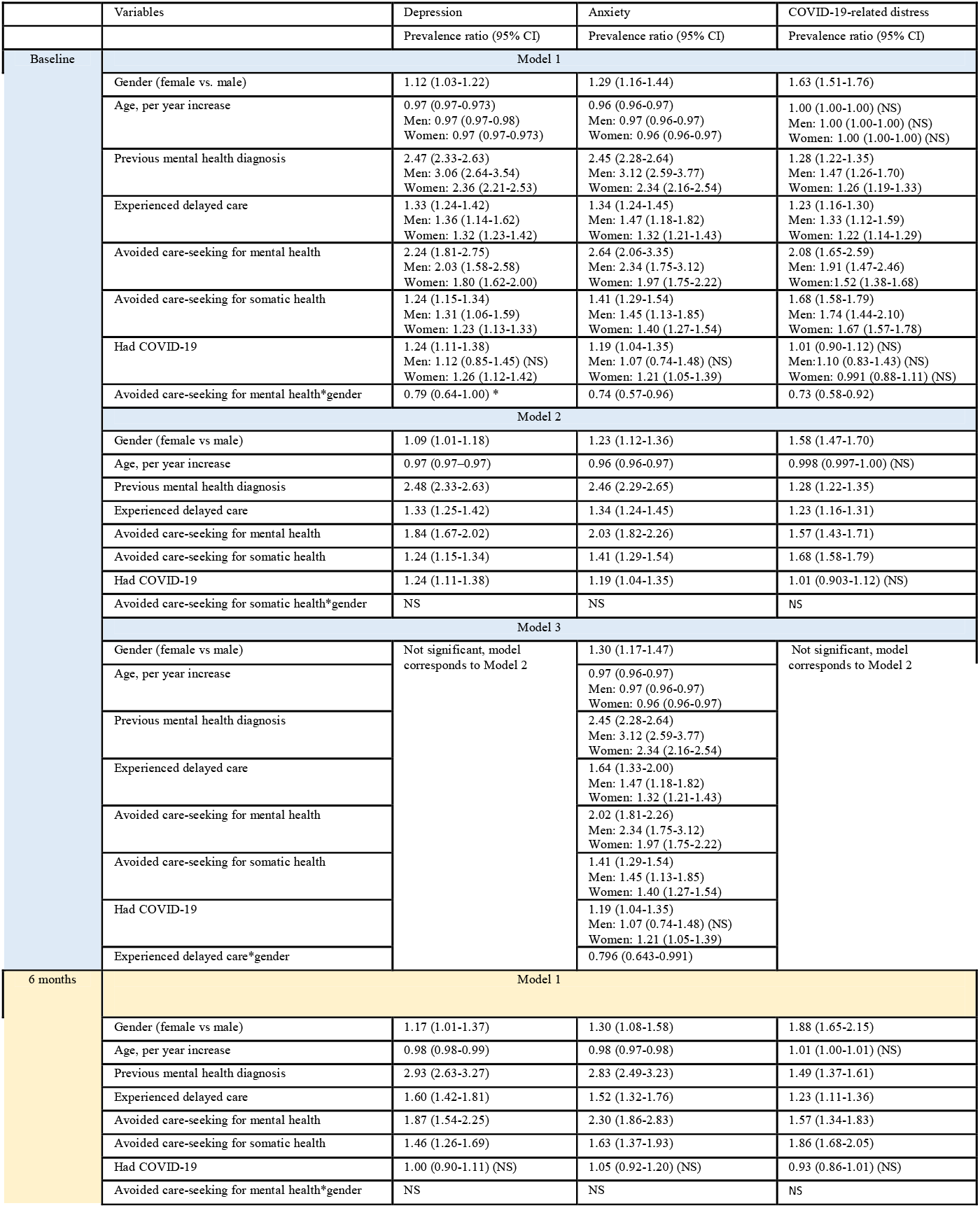
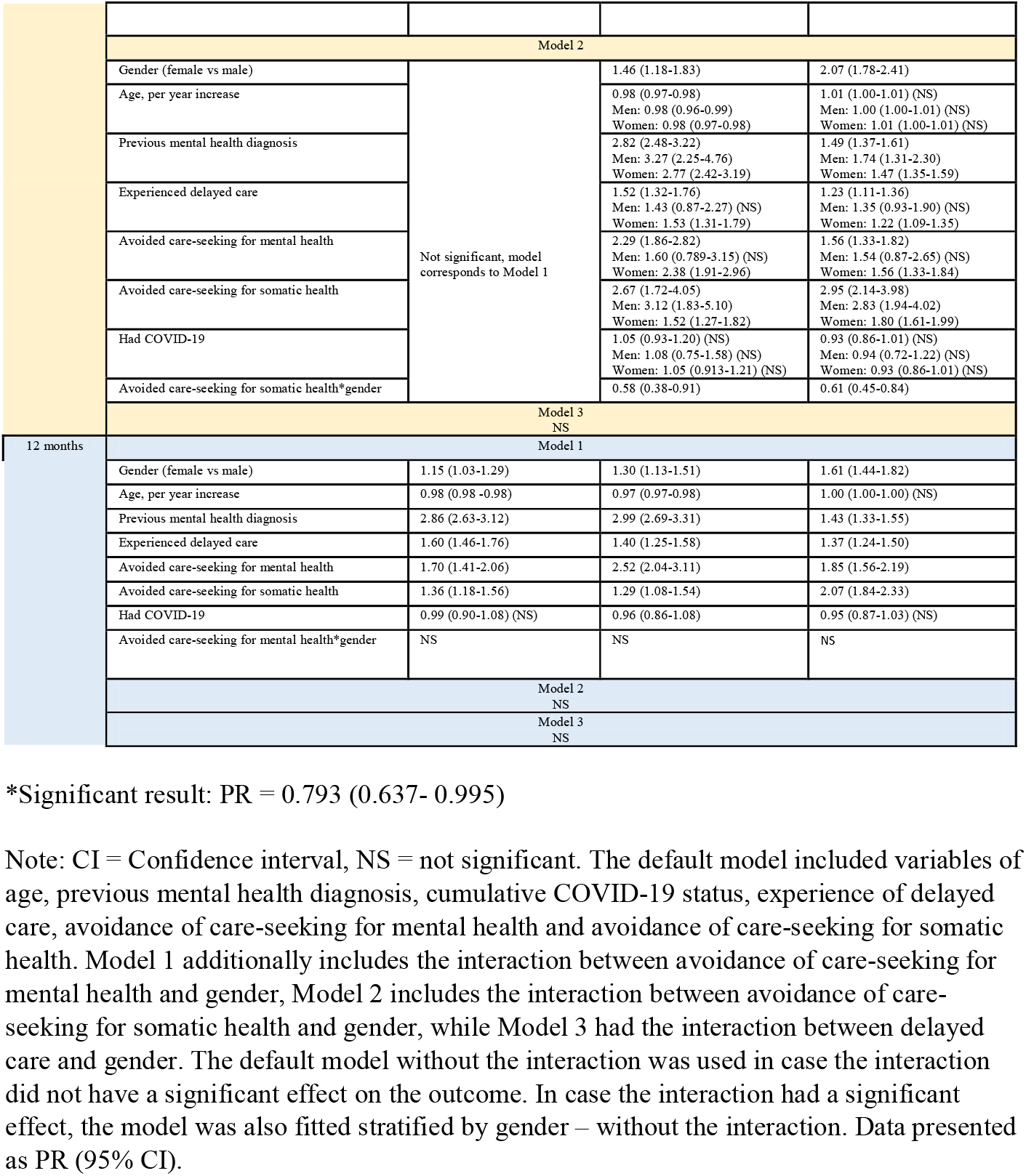
Prevalence ratios of depression, anxiety, and COVID-19-related distress in relation to gender and care-seeking and delayed care

## Discussion

In this study, we reported differences in mental health as well as care-seeking behavior between men and women over three time points during the COVID-19 pandemic in Sweden. We observed higher levels of depression, anxiety, and COVID-19-related distress as well as avoidance of care-seeking among women than men, as well as stronger associations of avoidance of care-seeking for mental or physical health with depression, anxiety, and COVID-19-related distress among men than women.

One of the important findings of this study is that women were more likely to report avoidance in seeking health care services for mental and somatic health problems. Prior to the COVID-19 pandemic, research suggested that men were more likely to avoid seeking care, while conflicting results have been found during the pandemic (4–7). In line with our findings, some studies found that women avoided seeking care more often (12,41) although other studies suggested that men utilized care less (13,20,42), during the pandemic. Nonetheless, a recent review indicates a general decrease in healthcare utilization for non-COVID-19 conditions almost universally, across genders, income levels, and countries, most likely due to a combination of factors, such as lockdown measures and fears of contracting the virus (43).

Network analysis indicated that female gender was positively associated with COVID-19-related distress and previous mental health diagnosis at baseline and 6-month follow-up, likely attributed to discussed gender difference in the prevalence of mental illness as well as diagnostics (2). Furthermore, female gender was positively associated with anxiety and avoidance of care-seeking for mental health due to worry of COVID-19 at 12-month follow-up. Regardless, anxiety was strongly positively correlated to depression at all time points, providing further evidence for the comorbidity of anxiety and depression and a high prevalence of these disorders among women (2,44,45).

Previous mental health diagnosis was shown as a predictor of depression, anxiety, and COVID-19-related distress, before and after stratification by gender. In the gender-stratified analyses, the association of previous mental health diagnosis with burden of mental health symptoms was stronger among men, compared to women. Similar pattern was noted for avoidance of care-seeking for mental health, avoidance of care-seeking for somatic health, as well as experiencing delayed care. This suggests that the impact of avoiding care-seeking and delayed care due to COVID-19 is greater among men than women, a finding that complements research on delayed and missed care leading to worse health outcomes (17,46,47). Finally, evidence has emerged suggesting a bi-directional relationship between poor mental health and a higher risk of COVID-19 (16,48), corroborating findings of the present study showing that previous COVID-19 diagnosis was associated with a higher risk of mental-ill health.

The strengths of this study lie in the 3-time point cross-sectional design that complements the existing evidence base on mental health during the pandemic in Sweden, which mainly consists of classical cross-sectional studies (25,49) and analysis of baseline data from longitudinal studies (20,26). Nevertheless, the data collection period for this study spanned over nearly two years and allows for comparison for changes over time. Limitations of our study must also be noted. Firstly, selection bias could have arisen from the voluntary nature of participation, meaning that the sample is unlikely to be representative of the general Swedish population. Furthermore, the low proportion of male participants (18.53%) limits our ability to draw robust conclusions regarding gender difference. The gender variable used in the study is also limited by the recognition of only two genders in the Swedish personal identification number. Additionally, the results of this study may have limited applicability to pandemic settings in other countries, as the Swedish mitigating approaches have often been interpreted as different, with more relaxed recommendations rather than strict policies (24,50). Furthermore, it was not possible to identify delays in mental health care specifically, as the question for type of delayed care did not have a specific option for mental healthcare. Finally, levels of actual care-seeking in this study could not be assessed, as data was only available on avoided or delayed care. Therefore, this represents an important area for future research.

## Conclusion

This study explored gender difference in mental health, care-seeking, as well as the association between care-seeking and mental health during the COVID-19 pandemic in Sweden. A discrepancy was observed between the genders in the prevalence of mental health symptoms as well as avoidance of care-seeking due to COVID-19. In the network analysis, at both baseline and 6-month follow-up, being a woman was positively associated with COVID-19-related distress and previous mental health diagnosis. At 12-month follow-up, female gender was additionally positively associated with anxiety and avoidance of mental health care-seeking. Regardless, a previous mental health diagnosis, avoidance of care-seeking, and experiencing delayed care were more strongly associated with negative mental health outcomes among men than women.

## Data Availability

All data produced in the present study are available upon reasonable request to the authors

## Acknowledgements

To all Omtanke2020 participants and researchers.

