## Supplements for "Mental health, gender, and care-seeking behavior during the COVID-19 pandemic in Sweden: An exploratory study"

**Supplementary materials**

- R code

\# LOAD DATA##################

library(readr)

data <- read_csv("data.csv")

View (data)

library(statip)

library(modest)

library(psych)

library(dplyr)

library (igraph)

library (ggraph)

library (networkD3)

library (writexl)

library (tidyverse)

library (corrr)

library (pastecs)

library (pcaPP)

library (bootnet)

library (summarytools)

library(igraph)

library (relaimpo)

library(tidyverse)

library(geepack)

library(table1)

library(emmeans)

library (mgm)

library(readxl)

library (mfx)

to_import <- read_excel("to import.xlsx")

View(to_import)

library (logbin)

library (epiR)

library (Epi)

library (foreign)

data <- merge (data, to_import, by= "userid", all.x = TRUE)

library(readxl)

to_import2 <- read_excel("to import2.xlsx")

View(to_import2)

data <- merge (data, to_import2, by= "userid", all.x = TRUE)

library(readxl)

to_import3 <- read_excel("to import3.xlsx")

View(to_import3)

data <- merge (data, to_import3, by= "userid", all.x= TRUE)

library(readxl)

to_import4 <- read_excel("to import4.xlsx")

View(to_import4)

data <- merge (data, to_import4, by= "userid", all.x= TRUE)

library(readxl)

ptsdbaseline <- read_excel("ptsdbaseline.xlsx")

View(ptsdbaseline)

data <- merge (data, ptsdbaseline, by= "userid", all.x= TRUE)

library(readxl)

ptsdhalfyear <- read_excel("ptsdhalfyear.xlsx")

View(ptsdhalfyear)

data <- merge (data, ptsdhalfyear, by= "userid", all.x= TRUE)

library(readxl)

ptsdoneyear <- read_excel("ptsdoneyear.xlsx")

View(ptsdoneyear)

data <- merge (data, ptsdoneyear, by= "userid", all.x= TRUE)

data$newcomorbsc <- recode(data$comorb_scale, "0"= 0, "1" =1, "2" =2, "3+" = 3)

data$ptsd0_1 <- recode(data$Q38_1, "1" = 0, "2" = 1, "3"= 1, "4"= 1, "5" = 1)

data$ptsd0_2 <- recode (data$Q38_2, "1" = 0, "2" = 1, "3"= 1, "4"= 1, "5" = 1)

data$ptsd0_3 <- recode (data$Q38_3, "1" = 0, "2" = 1, "3"= 1, "4"= 1, "5" = 1)

data$ptsd0_4 <- recode (data$Q38_4, "1" = 0, "2" = 1, "3"= 1, "4"= 1, "5" = 1)

data$ptsd0_5 <- recode (data$Q38_5, "1" = 0, "2" = 1, "3"= 1, "4"= 1, "5" = 1)

data$ptsd0 <- data$ptsd0_1 + data$ptsd0_2 + data$ptsd0_3 + data$ptsd0_4 + data$ptsd0_5

data$ptsd6_1 <- recode(data$m7_Q38_1, "1" = 0, "2" = 1, "3"= 1, "4"= 1, "5" = 1)

data$ptsd6_2 <- recode (data$m7_Q38_2, "1" = 0, "2" = 1, "3"= 1, "4"= 1, "5" = 1)

data$ptsd6_3 <- recode (data$m7_Q38_3, "1" = 0, "2" = 1, "3"= 1, "4"= 1, "5" = 1)

data$ptsd6_4 <- recode (data$m7_Q38_4, "1" = 0, "2" = 1, "3"= 1, "4"= 1, "5" = 1)

data$ptsd6_5 <- recode (data$m7_Q38_5, "1" = 0, "2" = 1, "3"= 1, "4"= 1, "5" = 1)

data$ptsd6 <- data$ptsd6_1 + data$ptsd6_2 + data$ptsd6_3 + data$ptsd6_4 + data$ptsd6_5

data$ptsd12_1 <- recode(data$m13_Q38_1, "1" = 0, "2" = 1, "3"= 1, "4"= 1, "5" = 1)

data$ptsd12_2 <- recode (data$m13_Q38_2, "1" = 0, "2" = 1, "3"= 1, "4"= 1, "5" = 1)

data$ptsd12_3 <- recode (data$m13_Q38_3, "1" = 0, "2" = 1, "3"= 1, "4"= 1, "5" = 1)

data$ptsd12_4 <- recode (data$m13_Q38_4, "1" = 0, "2" = 1, "3"= 1, "4"= 1, "5" = 1)

data$ptsd12_5 <- recode (data$m13_Q38_5, "1" = 0, "2" = 1, "3"= 1, "4"= 1, "5" = 1)

data$ptsd12 <- data$ptsd12_1 + data$ptsd12_2 + data$ptsd12_3 + data$ptsd12_4 + data$ptsd12_5

data$newFU0_Q35_4 <- recode(data$FU0_Q35_4, "1"= 1, "2" =0)

data$newFU6_Q35_4 <- recode(data$FU6_Q35_4, "1"= 1, "2" =0)

data$newFU12_Q35_4 <- recode(data$FU12_Q35_4, "1"= 1, "2" =0)

data$BMIcat <- "Underweight"

data[which(data$BMID == NA), 'BMIcat'] <- "Missing"

data[which(data$BMID>18.5 & data$BMID<24.99), 'BMIcat'] <- "Normal"

data[which(data$BMID>=25 & data$BMID<29.9), 'BMIcat'] <- "Overweight"

data[which(data$BMID>=30), 'BMIcat'] <- "Obese"

data$BMIcat <- as.factor(data$BMIcat)

data$PHQcat0 <- "Minimal"

data[which(data$FU0_total_score_PHQ9 == NA), 'PHQcat0'] <- "Missing"

data[which(data$FU0_total_score_PHQ9>=5 & data$FU0_total_score_PHQ9<10), 'PHQcat0'] <- "Mild"

data[which(data$FU0_total_score_PHQ9 >=10 & data$FU0_total_score_PHQ9<15), 'PHQcat0'] <- "Moderate"

data[which(data$FU0_total_score_PHQ9>=15 & data$FU0_total_score_PHQ9<20), 'PHQcat0'] <- "Moderately severe"

data[which(data$FU0_total_score_PHQ9>=20), 'PHQcat0'] <-"Severe"

data$PHQcat0 <- as.factor(data$PHQcat0)

data$PHQcat6 <- "Minimal"

data[which(data$FU6_total_score_PHQ9 == NA), 'PHQcat6'] <- "Missing"

data[which(data$FU6_total_score_PHQ9>=5 & data$FU6_total_score_PHQ9<10), 'PHQcat6'] <- "Mild"

data[which(data$FU6_total_score_PHQ9 >=10 & data$FU6_total_score_PHQ9<15), 'PHQcat6'] <- "Moderate"

data[which(data$FU6_total_score_PHQ9>=15 & data$FU6_total_score_PHQ9<20), 'PHQcat6'] <- "Moderately severe"

data[which(data$FU6_total_score_PHQ9>=20), 'PHQcat6'] <-"Severe"

data$PHQcat6 <- as.factor(data$PHQcat6)

data$PHQcat12 <- "Minimal"

data[which(data$FU12_total_score_PHQ9 == NA), 'PHQcat12'] <- "Missing"

data[which(data$FU12_total_score_PHQ9>=5 & data$FU12_total_score_PHQ9<10), 'PHQcat12'] <- "Mild"

data[which(data$FU12_total_score_PHQ9 >=10 & data$FU12_total_score_PHQ9<15), 'PHQcat12'] <- "Moderate"

data[which(data$FU12_total_score_PHQ9>=15 & data$FU12_total_score_PHQ9<20), 'PHQcat12'] <- "Moderately severe"

data[which(data$FU12_total_score_PHQ9>=20), 'PHQcat12'] <-"Severe"

data$PHQcat12 <- as.factor(data$PHQcat12)

data$GADcat0 <- "Minimal"

data[which(data$FU0_total_score_GAD7 == NA), 'GADcat0'] <- "Missing"

data[which(data$FU0_total_score_GAD7>=5 & data$FU0_total_score_GAD7<10), 'GADcat0'] <- "Mild"

data[which(data$FU0_total_score_GAD7>=10 & data$FU0_total_score_GAD7<15), 'GADcat0'] <- "Moderate"

data[which(data$FU0_total_score_GAD7>=15), 'GADcat0'] <- "Severe"

data$GADcat0 <- as.factor(data$GADcat0)

data$GADcat6 <- "Minimal"

data[which(data$FU6_total_score_GAD7 == NA), 'GADcat6'] <- "Missing"

data[which(data$FU6_total_score_GAD7>=5 & data$FU6_total_score_GAD7<10), 'GADcat6'] <- "Mild"

data[which(data$FU6_total_score_GAD7>=10 & data$FU6_total_score_GAD7<15), 'GADcat6'] <- "Moderate"

data[which(data$FU6_total_score_GAD7>=15), 'GADcat6'] <- "Severe"

data$GADcat6 <- as.factor(data$GADcat6)

data$GADcat12 <- "Minimal"

data[which(data$FU12_total_score_GAD7 == NA), 'GADcat12'] <- "Missing"

data[which(data$FU12_total_score_GAD7>=5 & data$FU12_total_score_GAD7<10), 'GADcat12'] <- "Mild"

data[which(data$FU12_total_score_GAD7>=10 & data$FU12_total_score_GAD7<15), 'GADcat12'] <- "Moderate"

data[which(data$FU12_total_score_GAD7>=15), 'GADcat12'] <- "Severe"

data$GADcat12 <- as.factor(data$GADcat12)

data$newsex <-

data[which(data$sex == 'Man'), 'newsex'] <- 0

data[which (data$sex == 'Kvinna'),'newsex'] <- 1

data$newsex <- as.numeric(data$newsex)

data$oroligscale <- data$FU0_Q35_43_1 + data$FU0_Q35_43_2 + data$FU0_Q35_43_3 + data$FU0_Q35_43_4 + data$FU0_Q35_43_5 + data$FU0_Q35_43_99

data$binaryPHQbl <-

data[which(data$PHQcat0 =="Minimal" ), 'binaryPHQbl'] <- 0

data[which(data$PHQcat0 =="Mild" ), 'binaryPHQbl'] <- 0

data[which(data$PHQcat0 == "Moderate" ), 'binaryPHQbl'] <- 1

data[which(data$PHQcat0 == "Moderately severe" ), 'binaryPHQbl'] <- 1

data[which(data$PHQcat0 == "Severe" ), 'binaryPHQbl'] <- 1

data$binaryPHQbl <- as.numeric (data$binaryPHQbl)

data$binaryPHQhalf <-

data[which(data$PHQcat6 =="Minimal" ), 'binaryPHQhalf'] <- 0

data[which(data$PHQcat6 =="Mild" ), 'binaryPHQhalf'] <- 0

data[which(data$PHQcat6 == "Moderate" ), 'binaryPHQhalf'] <- 1

data[which(data$PHQcat6 == "Moderately severe" ), 'binaryPHQhalf'] <- 1

data[which(data$PHQcat6 == "Severe" ), 'binaryPHQhalf'] <- 1

data$binaryPHQhalf <- as.numeric (data$binaryPHQhalf)

data$binaryPHQone <-

data[which(data$PHQcat12 =="Minimal" ), 'binaryPHQone'] <- 0

data[which(data$PHQcat12 =="Mild" ), 'binaryPHQone'] <- 0

data[which(data$PHQcat12 == "Moderate" ), 'binaryPHQone'] <- 1

data[which(data$PHQcat12 == "Moderately severe" ), 'binaryPHQone'] <- 1

data[which(data$PHQcat12 == "Severe" ), 'binaryPHQone'] <- 1

data$binaryPHQone <- as.numeric (data$binaryPHQone)

data$binaryGADbl <-

data[which(data$GADcat0 =="Minimal" ), 'binaryGADbl'] <- 0

data[which(data$GADcat0 =="Mild" ), 'binaryGADbl'] <- 0

data[which(data$GADcat0 == "Moderate" ), 'binaryGADbl'] <- 1

data[which(data$GADcat0 == "Severe" ), 'binaryGADbl'] <- 1

data$binaryGADbl <- as.numeric (data$binaryGADbl)

data$binaryGADhalf <-

data[which(data$GADcat6 =="Minimal" ), 'binaryGADhalf'] <- 0

data[which(data$GADcat6 =="Mild" ), 'binaryGADhalf'] <- 0

data[which(data$GADcat6 == "Moderate" ), 'binaryGADhalf'] <- 1

data[which(data$GADcat6 == "Severe" ), 'binaryGADhalf'] <- 1

data$binaryGADhalf <- as.numeric (data$binaryGADhalf)

data$binaryGADone <-

data[which(data$GADcat12 =="Minimal" ), 'binaryGADone'] <- 0

data[which(data$GADcat12 =="Mild" ), 'binaryGADone'] <- 0

data[which(data$GADcat12 == "Moderate" ), 'binaryGADone'] <- 1

data[which(data$GADcat12 == "Severe" ), 'binaryGADone'] <- 1

data$binaryGADone <- as.numeric (data$binaryGADone)

data$binaryptsd0 <-

data[which(data$ptsd0 == NA), 'binaryptsd0'] <- "Missing"

data[which(data$ptsd0 < 4), 'binaryptsd0'] <- 0

data[which(data$ptsd0 >= 4), 'binaryptsd0'] <- 1

data$binaryptsd0 <- as.numeric (data$binaryptsd0)

data$binaryptsd6 <-

data[which(data$ptsd6 == NA), 'binaryptsd6'] <- "Missing"

data[which(data$ptsd6 < 4), 'binaryptsd6'] <- 0

data[which(data$ptsd6 >= 4), 'binaryptsd6'] <- 1

data$binaryptsd6 <- as.numeric (data$binaryptsd6)

data$binaryptsd12 <-

data[which(data$ptsd12 == NA), 'binaryptsd12'] <- "Missing"

data[which(data$ptsd12 < 4), 'binaryptsd12'] <- 0

data[which(data$ptsd12 >= 4), 'binaryptsd12'] <- 1

data$binaryptsd12 <- as.numeric (data$binaryptsd12)

dfaa0 <- data.frame (data$prev_psy, data$newsex, data$age, data$comorb, data$FU0_total_score_PHQ9, data$ptsd0, data$FU0_total_score_GAD7, data$FU0_Q35_4,

data$FU0_Q35_2, data$FU0_Q35_3, data$FU0_covid_status)

print (dfaa0)

dfaa6 <- data.frame (data$prev_psy, data$newsex, data$age, data$comorb, data$FU6_total_score_PHQ9, data$ptsd6, data$FU6_total_score_GAD7, data$FU6_Q35_4,

data$FU6_Q35_2, data$FU6_Q35_3, data$FU6_covid_status)

print (dfaa6)

dfaa12 <- data.frame (data$prev_psy, data$newsex, data$age, data$comorb, data$FU12_total_score_PHQ9, data$ptsd12, data$FU12_total_score_GAD7, data$FU12_Q35_4,

data$FU12_Q35_2, data$FU12_Q35_3, data$FU12_covid_status)

print (dfaa12)

hist(sqrt)

hist (data$FU0_total_score_GAD7)

#this one is the problematic variable for RIN####

#data$FU0_Q35_4

df00 <- subset (data, data$FU0_Q35_4 == 1)

df0 <- data.frame (df00$prev_psy, df00$newsex, df00$age, df00$comorb, df00$FU0_Q35_2, df00$FU0_covid_status, df00$FU0_Q35_3, df00$FU0_Q35_43_single, df00$FU0_total_score_GAD7,

df00$ptsd0, df00$FU0_total_score_PHQ9)

print (df0)

df06 <- subset (data, data$FU6_Q35_4 ==1)

df6 <- data.frame (df06$prev_psy, df06$newsex, df06$age, df06$comorb, df06$FU6_covid_status, df06$FU6_total_score_PHQ9, df06$ptsd6, df06$FU6_total_score_GAD7,

df06$FU6_Q35_2,df06$FU6_Q35_3, df06$FU6_Q35_43_single)

print (df6)

df012 <- subset (data, data$FU12_Q35_4 == 1)

df12 <- data.frame (df012$prev_psy, df012$newsex, df012$age, df012$comorb, df012$FU12_covid_status, df012$FU12_total_score_PHQ9, df012$ptsd12, df012$FU12_total_score_GAD7,

df012$FU12_Q35_2, df012$FU12_Q35_3, df012$FU12_Q35_43_single)

print (df12)

dfmen <- subset (data, newsex == 0)

dfwomen <- subset (data, newsex == 1)

#DESCRIPTIVES#############

summary.data.frame(data)

summary(data$FU0_Q8)

sd(data$FU0_Q8, na.rm = TRUE)

mode = mfv(data$FU0_Q8)

print(mode)

hist(data$FU0_Q8[data$sex == "Kvinna"])

hist(data$FU0_Q8[data$sex == "Man"])

hist(data$FU0_Q9[data$sex == "Kvinna"])

hist(data$FU0_Q9[data$sex == "Man"])

summary (data$comorb_scale)

summary(data$newcomorbsc)

table (data$newcomorbsc)

hist (data$newcomorbsc)

table(data$comorb_scale)

table (data$`summa3 with average`)

hist (data$`summa3 with average`)

hist(data$newcomorbsc[data$sex == "Kvinna"])

hist(data$newcomorbsc[data$sex == "Man"])

summary(data$newcomorbsc[data$sex == "Kvinna"])

sd(data$newcomorbsc[data$sex == "Kvinna"], na.rm = TRUE)

summary(data$newcomorbsc[data$sex == "Man"])

sd(data$newcomorbsc[data$sex == "Man"], na.rm = TRUE)

summary(data$FU0_Q9)

sd(data$FU0_Q9, na.rm = TRUE)

mode = mfv(data$FU0_Q9)

print(mode)

summary (data$FU0_total_score_PCPTSD5)

sd(data$FU0_total_score_PCPTSD5, na.rm = TRUE)

summary (data$ptsd0)

sd(data$ptsd0, na.rm = TRUE)

summary (data$ptsd6)

sd(data$ptsd6, na.rm = TRUE)

summary (data$ptsd12)

sd(data$ptsd12, na.rm = TRUE)

summary (data$FU0_total_score_GAD7)

sd(data$FU0_total_score_GAD7, na.rm = TRUE)

summary (data$FU0_total_score_PSS4)

sd(data$FU0_total_score_PSS4, na.rm = TRUE)

summary (data$FU0_total_score_PHQ9)

sd(data$FU0_total_score_PHQ9, na.rm = TRUE)

summary (data$FU6_total_score_GAD7)

sd(data$FU6_total_score_GAD7, na.rm = TRUE)

summary (data$FU6_total_score_PHQ9)

sd(data$FU6_total_score_PHQ9, na.rm = TRUE)

summary (data$FU12_total_score_GAD7)

sd(data$FU12_total_score_GAD7, na.rm = TRUE)

summary (data$FU12_total_score_PHQ9)

sd(data$FU12_total_score_PHQ9, na.rm = TRUE)

summary (dfmen$age)summary (dfmen$FU0_total_score_PHQ9)

sd(dfmen$FU0_total_score_PHQ9, na.rm = TRUE)

summary (dfwomen$FU0_total_score_PHQ9)

sd(dfwomen$FU0_total_score_PHQ9, na.rm = TRUE)

summary (dfmen$FU0_total_score_GAD7)

sd(dfmen$FU0_total_score_GAD7, na.rm = TRUE)

summary (dfwomen$FU0_total_score_GAD7)

sd(dfwomen$FU0_total_score_GAD7, na.rm = TRUE)

summary (dfmen$ptsd0)

sd(dfmen$ptsd0, na.rm = TRUE)

summary (dfwomen$ptsd0)

sd(dfwomen$ptsd0, na.rm = TRUE)

summary (data$FU0_Q9)

sd (data$FU0_Q9, na.rm = TRUE)

summary (dfmen$FU0_Q9)

sd (dfmen$FU0_Q9, na.rm = TRUE)

summary (dfwomen$FU0_Q9)

sd (dfwomen$FU0_Q9, na.rm = TRUE)

summary (data$FU0_Q8)

sd (data$FU0_Q8, na.rm = TRUE)

summary (dfmen$FU0_Q8)

sd (dfmen$FU0_Q8, na.rm = TRUE)

summary (dfwomen$FU0_Q8)

sd (dfwomen$FU0_Q8, na.rm = TRUE)

summary (dfmen$age)

sd(dfmen$age, na.rm = TRUE)

summary (dfwomen$age)

sd(dfwomen$age, na.rm = TRUE)

summary (data$age)

sd(data$age, na.rm = TRUE)

describeBy(data$FU0_Q8, group = data$newsex)

describeBy(data$FU0_Q9, group = data$newsex)

table (data$FU0_Q43_1)

table (data$FU0_Q43_2)

table (data$FU0_Q43_3)

table (data$FU0_Q43_4)

table (data$FU0_Q43_5)

table (data$FU0_Q43_6)

table (data$FU0_Q43_7)

table(data$FU0_covid_status)

table (data$FU6_covid_status)

table (data$FU12_covid_status)

table(data$sex)

table(data$relation)

table(data$prev_psy)

table (data$binge_drink)

table (data$comorb)

table(data$comorb_scale)

table (data$self_rec)

table (data$smoking)

table (data$age_range)

table (data$FU0_bed)

table (data$FU6_bed)

table (data$FU12_bed)

table (data$FU0_Q8)

table (data$FU0_Q9)

table (data$FU0_Q35_2)

table (data$FU0_Q35_3)

table (data$FU0_Q35_4)

table (data$FU0_Q35_41)

table(data$FU0_Q35_42_1)

table(data$FU0_Q35_42_2)

table(data$FU0_Q35_42_3)

table(data$FU0_Q35_42_4)

table (data$Q35_42_5)

table (data$FU0_Q35_43_single)

table (data$FU6_Q8)

table (data$FU6_Q9)

table (data$FU6_Q35_2)

table (data$FU6_Q35_3)

table (data$FU6_Q35_4)

table (data$FU6_Q35_41)

table (data$FU6_Q35_43_single)

table (data$FU12_Q8)

table (data$FU12_Q9)

table (data$FU12_Q35_2)

table (data$FU12_Q35_3)

table (data$FU12_Q35_4)

table (data$FU12_Q35_41)

table (data$FU12_Q35_43_single)

table (data$FU0_Q35_42_1)

table (data$FU0_Q35_42_2)

table (data$FU0_Q35_42_3)

table (data$FU0_Q35_42_4)

table (data$Q35_42_5)

table (data$FU6_Q35_42_1)

table (data$FU6_Q35_42_2)

table (data$FU6_Q35_42_3)

table (data$FU6_Q35_42_4)

table (data$m7_Q35_42_5)

table (data$FU12_Q35_42_1)

table (data$FU12_Q35_42_2)

table (data$FU12_Q35_42_3)

table (data$FU12_Q35_42_4)

table (data$m13_Q35_42_5)

table (data$FU0_Q35_43_1)

table (data$FU0_Q35_43_2)

table (data$FU0_Q35_43_3)

table (data$FU0_Q35_43_4)

table (data$FU0_Q35_43_5)

table (data$FU0_Q35_43_99)

#comorbidities:

table (data$FU0_Q43_1)

table (data$FU0_Q43_2)

table (data$FU0_Q43_3)

table (data$FU0_Q43_4)

table (data$FU0_Q43_5)

table (data$FU0_Q43_6)

table (data$FU0_Q43_7)

sd(data$age)

plot(data$BMID)

hist (data$age)

hist (data$smoking)

#BASELINE MENTAL HEALTH###########

hist (data$FU0_total_score_GAD7,

breaks = 7,

main = "Histogram of anxiety scores (baseline)",

ylab = "frequency",

xlab = "score on GAD7"

)

hist (data$FU0_total_score_PHQ9,

breaks = 10,

main = "Histogram of depression scores (baseline)",

ylab = "frequency",

xlab = "score on PHQ9"

)

hist (data$FU0_total_score_PCPTSD5,

breaks = 5,

main = "Histogram of PTSD values (baseline)",

ylab = "frequency",

xlab = "score on PCPTSD5"

)

hist (data$ptsd0,

breaks = 5,

main = "Histogram of PTSD values (baseline)",

ylab = "frequency",

xlab = "score on PCPTSD5"

)

hist (data$FU0_total_score_PSS4,

breaks = 4,

main = "Histogram of stress scores (baseline)",

ylab = "frequency",

xlab = "score on PSS4"

)

#FIRST FOLLOW-UP MENTAL HEALTH####

hist (data$FU6_total_score_GAD7,

breaks = 7,

main = "Histogram of anxiety scores (first follow-up)",

ylab = "frequency",

xlab = "score on GAD7"

)

hist (data$FU6_total_score_PHQ9,

breaks = 10,

main = "Histogram of depression scores (first follow-up)",

ylab = "frequency",

xlab = "score on PHQ9"

)

hist (data$FU6_total_score_PCPTSD5,

breaks = 5,

main = "Histogram of PTSD values (first follow-up)",

ylab = "frequency",

xlab = "score on PCPTSD5"

)

hist (data$ptsd6,

breaks = 5,

main = "Histogram of PTSD values (first follow-up)",

ylab = "frequency",

xlab = "score on PCPTSD5"

)

hist (data$FU6_total_score_PSS4,

breaks = 4,

main = "Histogram of stress scores (first follow-up)",

ylab = "frequency",

xlab = "score on PSS4"

)

#SECOND FOLLOW-UP MENTAL HEALTH##

hist (data$FU12_total_score_GAD7,

breaks = 7,

main = "Histogram of anxiety scores (second follow-up)",

ylab = "frequency",

xlab = "score on GAD7"

)

hist (data$FU12_total_score_PHQ9,

breaks = 10,

main = "Histogram of depression scores (second follow-up)",

ylab = "frequency",

xlab = "score on PHQ9"

)

hist (data$FU12_total_score_PCPTSD5,

breaks = 5,

main = "Histogram of PTSD values (second follow-up)",

ylab = "frequency",

xlab = "score on PCPTSD5"

)

hist (data$ptsd12,

breaks = 5,

main = "Histogram of PTSD values (second follow-up)",

ylab = "frequency",

xlab = "score on PCPTSD5"

)

hist (data$FU12_total_score_PSS4,

breaks = 4,

main = "Histogram of stress scores (second follow-up)",

ylab = "frequency",

xlab = "score on PSS4"

)

#CONTINGENCY TABLES########

Table = table(data$sex, data$age_range)

print(Table)

Table = table (data$PHQcat6, data$sex)

print(Table)

Table = table(data$sex, data$relation)

print (Table)

Table = table (data$sex, data$relation, useNA = 'always')

print (Table)

Table = table (data$sex, data$BMIcat)

print (Table)

Table = table (data$sex, data$PHQcat0)

print (Table)

Table = table (data$sex, data$FU0_total_score_PHQ9)

print (Table)

Table = table (data$sex, data$ptsd0)

print (Table)

table (data$ptsd0)

Table = table (data$sex, data$GADcat0)

print (Table)

Table = table (data$sex, data$binaryptsd0)

print (Table)

table(is.na(data$BMID), data$sex)

Table = table (data$sex, data$smoking)

print (Table)

table(is.na(data$smoking), data$sex)

Table = table (data$sex, data$binge_drink)

print (Table)

table(is.na(data$binge_drink), data$sex)

Table = table (data$sex, data$`summa3 with average`)

print (Table)

table(is.na(data$`summa3 with average`), data$sex)

Table = table (data$sex, data$comorb_scale)

print (Table)

table(is.na(data$comorb_scale), data$sex)

Table = table (data$sex, data$prev_psy)

print (Table)

table(is.na(data$prev_psy), data$sex)

Table = table (data$sex, data$FU0_covid_status)

print (Table)

table (is.na(data$FU0_covid_status), data$sex)

Table = table (data$sex, data$FU6_covid_status)

print (Table)

table (is.na(data$FU6_covid_status), data$sex)

Table = table (data$sex, data$FU12_covid_status)

print (Table)

table (is.na(data$FU12_covid_status), data$sex)

Table = table (data$sex, data$self_rec)

print (Table)

table(is.na(data$self_rec), data$sex)

Table= table(data$FU0_Q8, data$sex)

print (Table)

table(is.na(data$FU0_Q8), data$sex)

Table= table(data$FU0_Q9, data$sex)

print (Table)

table(is.na(data$FU0_Q9), data$sex)

Table = table(data$sex, data$FU0_Q35_2)

print(Table)

Table = table(data$sex, data$FU0_Q35_3)

print (Table)

Table = table(data$sex, data$FU0_Q35_4)

print (Table)

Table = table(data$sex, data$FU0_Q35_41)

print (Table)

Table = table(data$sex, data$FU0_Q35_42_1)

print (Table)

Table = table(data$sex, data$FU0_Q35_42_2)

print (Table)

Table = table(data$sex, data$FU0_Q35_42_3)

print (Table)

Table = table(data$sex, data$FU0_Q35_42_4)

print (Table)

Table = table(data$sex, data$Q35_42_5)

print (Table)

Table = table (data$sex, data$FU6_Q35_42_1)

print (Table)

Table = table (data$sex, data$FU6_Q35_42_2)

print (Table)

Table= table (data$sex, data$FU6_Q35_42_3)

print (Table)

Table = table (data$sex, data$FU6_Q35_42_4)

print (Table)

Table = table (data$sex, data$m7_Q35_42_5)

print (Table)

Table = table (data$sex, data$FU12_Q35_42_1)

print (Table)

Table = table (data$sex, data$FU12_Q35_42_2)

print (Table)

Table = table (data$sex, data$FU12_Q35_42_3)

print (Table)

Table= table (data$sex, data$FU12_Q35_42_4)

print (Table)

Table = table (data$sex, data$m13_Q35_42_5)

print (Table)

Table = table (data$sex, data$FU0_Q43_1)

print (Table)

Table = table (data$sex, data$FU0_Q43_2)

print (Table)

Table = table(data$sex, data$FU0_Q43_3)

print (Table)

Table = table (data$sex, data$FU0_Q43_4)

print (Table)

Table= table (data$sex, data$FU0_Q43_5)

print (Table)

Table = table (data$sex, data$FU0_Q43_6)

print (Table)

Table = table (data$sex, data$FU0_Q43_7)

print (Table)

table(is.na(data$FU0_Q43_1), data$sex)

table(is.na(data$FU0_Q43_2), data$sex)

table(is.na(data$FU0_Q43_3), data$sex)

table(is.na(data$FU0_Q43_4), data$sex)

table(is.na(data$FU0_Q43_5), data$sex)

table(is.na(data$FU0_Q43_6), data$sex)

table(is.na(data$FU0_Q43_7), data$sex)

table(is.na(data$FU0_Q35_2), data$sex)

table(is.na(data$FU0_Q35_3), data$sex)

table(is.na(data$FU0_Q35_4), data$sex)

table(is.na(data$FU0_Q35_41), data$sex)

table(is.na(data$FU0_Q35_42_1), data$sex)

table(is.na(data$FU0_Q35_42_3), data$sex)

table(is.na(data$FU0_Q35_42_4), data$sex)

table(is.na(data$FU0_Q35_43_single), data$sex)

table(is.na(data$FU6_Q35_2), data$sex)

table(is.na(data$FU6_Q35_3), data$sex)

table(is.na(data$FU6_Q35_4), data$sex)

table(is.na(data$FU6_Q35_41), data$sex)

table(is.na(data$FU6_Q35_42_4), data$sex)

table(is.na(data$FU6_Q35_43_single), data$sex)

table(is.na(data$FU12_Q35_2), data$sex)

table(is.na(data$FU12_Q35_3), data$sex)

table(is.na(data$FU12_Q35_4), data$sex)

table(is.na(data$FU12_Q35_41), data$sex)

table(is.na(data$FU12_Q35_42_2), data$sex)

table(is.na(data$m13_Q35_42_5), data$sex)

table(is.na(data$FU12_Q35_43_single), data$sex)

table(is.na(dfmen$ptsd0))

table(is.na(dfwomen$ptsd0))

#MAYBE THESE DO NOT MAKE ANY SENSE###

Table = table (data$sex, data$FU0_Q35_43_1)

print (Table)

Table = table (data$sex, data$FU0_Q35_43_2)

print (Table)

Table = table (data$sex, data$FU0_Q35_43_3)

print (Table)

Table = table (data$sex, data$FU0_Q35_43_4)

print (Table)

Table = table (data$sex, data$FU0_Q35_43_5)

print (Table)

Table = table (data$sex, data$FU0_Q35_43_99)

print (Table)

table(is.na(data$FU0_Q35_43_1), data$sex)

table(is.na(data$FU0_Q35_43_2), data$sex)

table(is.na(data$FU0_Q35_43_3), data$sex)

table(is.na(data$FU0_Q35_43_4), data$sex)

table(is.na(data$FU0_Q35_43_5), data$sex)

table(is.na(data$FU0_Q35_43_99), data$sex)

\###

Table = table (data$sex, data$FU0_Q35_43_single)

print (Table)

table(is.na(data$FU0_Q35_43_single), data$sex)

Table = table(data$sex, data$FU6_Q35_2)

print(Table)

Table = table(data$sex, data$FU6_Q35_3)

print (Table)

Table = table(data$sex, data$FU6_Q35_4)

print (Table)

Table = table(data$sex, data$FU6_Q35_41)

print (Table)

Table = table (data$sex, data$FU6_Q35_43_single)

print (Table)

Table = table(data$sex, data$FU12_Q35_2)

print(Table)

Table = table(data$sex, data$FU12_Q35_3)

print (Table)

Table = table(data$sex, data$FU12_Q35_4)

print (Table)

Table = table(data$sex, data$FU12_Q35_41)

print (Table)

Table = table (data$sex, data$FU12_Q35_43_single)

print (Table)

table (data$BMIcat)

table (data$PHQcat0)

table (data$GADcat0)

table (data$GADcat0, data$prev_psy)

table(data$comorb, data$GADcat0)

counts <- table(data$sex, data$FU0_total_score_PHQ9)

barplot(counts, main="PH9 Distribution by sex",

xlab="PHQ9 score", col=c("red", "darkblue"),

legend = rownames(counts))

table (data$newsex)

table(data$oroligscale)

sum(data$FU0_Q35_43_1)

#MISSING#########

table(is.na(data$sex))

table(is.na(data$age_range))

table(is.na(data$relation))

table(is.na(data$BMID))

table(is.na(data$smoking))

table(is.na(data$binge_drink))

table(is.na(data$comorb_scale))

table (is.na(data$comorb))

table(is.na(data$prev_psy))

table(is.na(data$self_rec))

table(is.na(data$FU0_Q35_43_single))

table (is.na(data$FU0_Q8))

table (is.na(data$FU0_Q9))

table (is.na(data$FU0_Q35_42_1))

##and so on

table (is.na(data$FU0_Q43_1))

table (is.na(data$FU0_Q43_2))

table (is.na(data$FU0_Q43_3))

table (is.na(data$FU0_Q43_4))

table (is.na(data$FU0_Q43_5))

table (is.na(data$FU0_Q43_6))

table (is.na(data$FU0_Q43_7))

table (is.na(data$FU0_covid_status))

table (is.na(data$FU6_covid_status))

table (is.na(data$FU12_covid_status))

table(is.na(data$FU0_total_score_GAD7))

table(is.na(data$FU0_total_score_PCPTSD5))

table(is.na(data$ptsd0))

table (is.na(data$FU0_total_score_PHQ9))

table (is.na(data$FU0_total_score_PSS4))

table (is.na(data$FU0_Q35_2))

table (is.na(data$FU0_Q35_3))

table (is.na(data$FU0_Q35_4))

table (is.na(data$FU0_Q35_41))

table (is.na(data$FU6_Q35_2))

table (is.na(data$FU6_Q35_3))

table (is.na(data$FU6_Q35_4))

table (is.na(data$FU6_Q35_41))

table(is.na(data$m7_Q35_42_5))

table(is.na(data$FU6_Q35_42_4))

table(is.na(data$FU6_Q35_43_single))

table(is.na(data$FU12_Q35_2))

table(is.na(data$FU12_Q35_3))

table(is.na(data$FU12_Q35_4))

table(is.na(data$FU12_Q35_41))

table(is.na(data$FU0_total_score_PHQ9))

table(is.na(data$FU6_total_score_PHQ9))

table(is.na(data$FU12_total_score_PHQ9))

table(is.na(data$FU6_total_score_PHQ9), data$sex)

table(is.na(data$FU12_total_score_PHQ9), data$sex)

table(is.na(data$FU0_total_score_GAD7))

table(is.na(data$FU6_total_score_GAD7))

table(is.na(data$FU12_total_score_GAD7))

table(is.na(data$FU6_total_score_GAD7), data$sex)

table(is.na(data$FU12_total_score_GAD7), data$sex)

table(is.na(data$ptsd0))

table(is.na(data$ptsd6))

table(is.na(data$ptsd12))

table(is.na(data$FU12_Q35_42_4))

table(is.na(data$m13_Q35_42_5))

table (is.na(data$FU12_Q35_43_single))

table (is.na(data$`summa3 with average`))

as.numeric (data$newsex)

summary (dfmen)

\# MGM#####

colnames(dfaa0)[colnames(dfaa0) == "data.age"] = "Age"

colnames(dfaa0)[colnames(dfaa0) == "data.FU0_Q35_4"] = "Delay in care"

colnames(dfaa0)[colnames(dfaa0) == "data.newsex"] = "Gender"

colnames(dfaa0)[colnames(dfaa0) == "data.FU0_total_score_GAD7"] = "Anxiety"

colnames(dfaa0)[colnames(dfaa0) == "data.ptsd0"] = "Distress"

colnames(dfaa0)[colnames(dfaa0) == "data.FU0_total_score_PHQ9"] = "Depression"

colnames(dfaa0)[colnames(dfaa0) == "data.prev_psy"] = "MH diagnosis"

colnames(dfaa0)[colnames(dfaa0) == "data.comorb"] = "Comorbidity"

colnames(dfaa0)[colnames(dfaa0) == "data.FU0_Q35_2"] ="Avoid MH care"

colnames(dfaa0)[colnames(dfaa0) == "data.FU0_covid_status"] = "COVID-19"

colnames(dfaa0)[colnames(dfaa0) == "data.FU0_Q35_3"] = "Avoid SH care"

jpeg("FIGURE_00.jpg", units="in",height=5, width=3, res=600)

Time00 <- estimateNetwork(dfaa0, default = "mgm", Method = "cor_auto", nboot=10000)

plot(Time00, layout = 'spring', legend = F, title = "MGM Baseline", vsize =15)

dev.off()

colnames(dfaa6)[colnames(dfaa6) == "data.age"] = "Age"

colnames(dfaa6)[colnames(dfaa6) == "data.FU6_Q35_4"] = "Delay in care"

colnames(dfaa6)[colnames(dfaa6) == "data.newsex"] = "Gender"

colnames(dfaa6)[colnames(dfaa6) == "data.FU6_total_score_GAD7"] = "Anxiety"

colnames(dfaa6)[colnames(dfaa6) == "data.ptsd6"] = "Distress"

colnames(dfaa6)[colnames(dfaa6) == "data.FU6_total_score_PHQ9"] = "Depression"

colnames(dfaa6)[colnames(dfaa6) == "data.prev_psy"] = "MH diagnosis"

colnames(dfaa6)[colnames(dfaa6) == "data.comorb"] = "Comorbidity"

colnames(dfaa6)[colnames(dfaa6) == "data.FU6_Q35_2"] ="Avoid MH care"

colnames(dfaa6)[colnames(dfaa6) == "data.FU6_covid_status"] = "COVID-19"

colnames(dfaa6)[colnames(dfaa6) == "data.FU6_Q35_3"] = "Avoid SH care"

jpeg("FIGURE_06.jpg", units="in",height=5, width=3, res=600)

Time0half <- estimateNetwork(dfaa6, default = "mgm", Method = "cor_auto", nboot=10000)

plot(Time0half, layout = 'spring', legend = F, title = "MGM 6 months", vsize =15)

dev.off()

colnames(dfaa12)[colnames(dfaa12) == "data.age"] = "Age"

colnames(dfaa12)[colnames(dfaa12) == "data.FU12_Q35_4"] = "Delay in care"

colnames(dfaa12)[colnames(dfaa12) == "data.newsex"] = "Gender"

colnames(dfaa12)[colnames(dfaa12) == "data.FU12_total_score_GAD7"] = "Anxiety"

colnames(dfaa12)[colnames(dfaa12) == "data.ptsd12"] = "Distress"

colnames(dfaa12)[colnames(dfaa12) == "data.FU12_total_score_PHQ9"] = "Depression"

colnames(dfaa12)[colnames(dfaa12) == "data.prev_psy"] = "MH diagnosis"

colnames(dfaa12)[colnames(dfaa12) == "data.comorb"] = "Comorbidity"

colnames(dfaa12)[colnames(dfaa12) == "data.FU12_Q35_2"] ="Avoid MH care"

colnames(dfaa12)[colnames(dfaa12) == "data.FU12_covid_status"] = "COVID-19"

colnames(dfaa12)[colnames(dfaa12) == "data.FU12_Q35_3"] = "Avoid SH care"

jpeg("FIGURE_012.jpg", units="in",height=5, width=3, res=600)

Time0year <- estimateNetwork(dfaa12, default = "mgm", Method = "cor_auto", nboot=10000)

plot(Time0year, layout = 'spring', legend = F, title = "MGM 12 months", vsize =15)

dev.off()

intPHQ01 <- glm (binaryPHQbl ~ newsex + age + prev_psy + newFU0_Q35_4 + FU0_Q35_2 + FU0_Q35_3 + FU0_covid_status + FU0_Q35_2 * newsex, data = data,

family = poisson (link = log))

intPHQ01

coefs <- tidy(intPHQ01, exponentiate = TRUE, conf.int = TRUE)

coefs

menPHQ01 <- glm (binaryPHQbl ~ age + prev_psy + newFU0_Q35_4 + FU0_Q35_2 + FU0_Q35_3 + FU0_covid_status, data = dfmen,

family = poisson (link = log))

menPHQ01

coefs<- tidy(menPHQ01, exponentiate = TRUE, conf.int = TRUE)

coefs

womenPHQ01 <- glm (binaryPHQbl ~ age + prev_psy + newFU0_Q35_4 + FU0_Q35_2 + FU0_Q35_3 + FU0_covid_status, data = dfwomen,

family = poisson (link = log))

womenPHQ01

coefs<- tidy(womenPHQ01, exponentiate = TRUE, conf.int = TRUE)

coefs

intPHQ02 <- glm (binaryPHQbl ~ newsex + age + prev_psy + newFU0_Q35_4 + FU0_Q35_2 + FU0_Q35_3 + FU0_covid_status + FU0_Q35_3 * newsex, data = data,

family = poisson (link = log))

intPHQ02

coefs <- tidy(intPHQ02, exponentiate = TRUE, conf.int = TRUE)

coefs #-> not significant, so#

intPHQ02minus <- glm (binaryPHQbl ~ newsex + age + prev_psy + newFU0_Q35_4 + FU0_Q35_2 + FU0_Q35_3 + FU0_covid_status, data = data,

family = poisson (link = log))

intPHQ02minus

coefs <- tidy(intPHQ02minus, exponentiate = TRUE, conf.int = TRUE)

coefs

intPHQ03 <- glm (binaryPHQbl ~ newsex + age + prev_psy + newFU0_Q35_4 + FU0_Q35_2 + FU0_Q35_3 + FU0_covid_status + newFU0_Q35_4 * newsex, data = data,

family = poisson (link = log))

intPHQ03

coefs <- tidy(intPHQ03, exponentiate = TRUE, conf.int = TRUE)

coefs #-> not significant, so#

intPHQ03minus <- glm (binaryPHQbl ~ newsex + age + prev_psy + newFU0_Q35_4 + FU0_Q35_2 + FU0_Q35_3 + FU0_covid_status, data = data,

family = poisson (link = log))

intPHQ03minus

coefs <- tidy(intPHQ03minus, exponentiate = TRUE, conf.int = TRUE)

coefs

intPHQ61 <- glm (binaryPHQhalf ~ newsex + age + prev_psy + newFU6_Q35_4 + FU6_Q35_2 + FU6_Q35_3 + FU6_covid_status + FU6_Q35_2 * newsex, data = data,

family = poisson (link = log))

intPHQ61

coefs <- tidy(intPHQ61, exponentiate = TRUE, conf.int = TRUE)

coefs #-> not significant, so#

intPHQ61minus <- glm (binaryPHQhalf ~ newsex + age + prev_psy + newFU6_Q35_4 + FU6_Q35_2 + FU6_Q35_3 + FU6_covid_status, data = data,

family = poisson (link = log))

intPHQ61minus

coefs <- tidy(intPHQ61minus, exponentiate = TRUE, conf.int = TRUE)

coefs

intPHQ62 <- glm (binaryPHQhalf ~ newsex + age + prev_psy + newFU6_Q35_4 + FU6_Q35_2 + FU6_Q35_3 + FU6_covid_status + FU6_Q35_3 * newsex, data = data,

family = poisson (link = log))

intPHQ62

coefs <- tidy(intPHQ62, exponentiate = TRUE, conf.int = TRUE)

coefs#-> not significant, so#

intPHQ62minus <- glm (binaryPHQhalf ~ newsex + age + prev_psy + newFU6_Q35_4 + FU6_Q35_2 + FU6_Q35_3 + FU6_covid_status, data = data,

family = poisson (link = log))

intPHQ62minus

coefs <- tidy(intPHQ62minus, exponentiate = TRUE, conf.int = TRUE)

coefs

intPHQ63 <- glm (binaryPHQhalf ~ newsex + age + prev_psy + newFU6_Q35_4 + FU6_Q35_2 + FU6_Q35_3 + FU6_covid_status + newFU6_Q35_4 * newsex, data = data,

family = poisson (link = log))

intPHQ63

coefs <- tidy(intPHQ63, exponentiate = TRUE, conf.int = TRUE)

coefs#-> not significant, so#

intPHQ63minus <- glm (binaryPHQhalf ~ newsex + age + prev_psy + newFU6_Q35_4 + FU6_Q35_2 + FU6_Q35_3 + FU6_covid_status, data = data,

family = poisson (link = log))

intPHQ63minus

coefs <- tidy(intPHQ63minus, exponentiate = TRUE, conf.int = TRUE)

coefs

intPHQ121 <- glm (binaryPHQone ~ newsex + age + prev_psy + newFU12_Q35_4 + FU12_Q35_2 + FU12_Q35_3 + FU12_covid_status + FU12_Q35_2 * newsex, data = data,

family = poisson (link = log))

intPHQ121

coefs <- tidy(intPHQ121, exponentiate = TRUE, conf.int = TRUE)

coefs#-> not significant, so#

intPHQ121minus <- glm (binaryPHQone ~ newsex + age + prev_psy + newFU12_Q35_4 + FU12_Q35_2 + FU12_Q35_3 + FU12_covid_status, data = data,

family = poisson (link = log))

intPHQ121minus

coefs <- tidy(intPHQ121minus, exponentiate = TRUE, conf.int = TRUE)

coefs

intPHQ122 <- glm (binaryPHQone ~ newsex + age + prev_psy + newFU12_Q35_4 + FU12_Q35_2 + FU12_Q35_3 + FU12_covid_status + FU12_Q35_3 * newsex, data = data,

family = poisson (link = log))

intPHQ122

coefs <- tidy(intPHQ122, exponentiate = TRUE, conf.int = TRUE)

coefs#-> not significant, so#

intPHQ122minus <- glm (binaryPHQone ~ newsex + age + prev_psy + newFU12_Q35_4 + FU12_Q35_2 + FU12_Q35_3 + FU12_covid_status, data = data,

family = poisson (link = log))

intPHQ122minus

coefs <- tidy(intPHQ122minus, exponentiate = TRUE, conf.int = TRUE)

coefs

intPHQ123 <- glm (binaryPHQone ~ newsex + age + prev_psy + newFU12_Q35_4 + FU12_Q35_2 + FU12_Q35_3 + FU12_covid_status + newFU12_Q35_4 * newsex, data = data,

family = poisson (link = log))

intPHQ123

coefs <- tidy(intPHQ123, exponentiate = TRUE, conf.int = TRUE)

coefs#-> not significant, so#

intPHQ123minus <- glm (binaryPHQone ~ newsex + age + prev_psy + newFU12_Q35_4 + FU12_Q35_2 + FU12_Q35_3 + FU12_covid_status, data = data,

family = poisson (link = log))

intPHQ123minus

coefs <- tidy(intPHQ123minus, exponentiate = TRUE, conf.int = TRUE)

coefs

intGAD01 <- glm (binaryGADbl ~ newsex + age + prev_psy + newFU0_Q35_4 + FU0_Q35_2 + FU0_Q35_3 + FU0_covid_status + FU0_Q35_2 * newsex, data = data,

family = poisson (link = log))

intGAD01

coefs <- tidy(intGAD01, exponentiate = TRUE, conf.int = TRUE)

coefs

menGAD01 <- glm (binaryGADbl ~ age + prev_psy + newFU0_Q35_4 + FU0_Q35_2 + FU0_Q35_3 + FU0_covid_status, data = dfmen,

family = poisson (link = log))

menGAD01

coefs<- tidy(menGAD01, exponentiate = TRUE, conf.int = TRUE)

coefs

womenGAD01 <- glm (binaryGADbl ~ age + prev_psy + newFU0_Q35_4 + FU0_Q35_2 + FU0_Q35_3 + FU0_covid_status, data = dfwomen,

family = poisson (link = log))

womenGAD01

coefs<- tidy(womenGAD01, exponentiate = TRUE, conf.int = TRUE)

coefs

intGAD02 <- glm (binaryGADbl ~ newsex + age + prev_psy + newFU0_Q35_4 + FU0_Q35_2 + FU0_Q35_3 + FU0_covid_status + FU0_Q35_3 * newsex, data = data,

family = poisson (link = log))

intGAD02

coefs <- tidy(intGAD02, exponentiate = TRUE, conf.int = TRUE)

coefs #-> not significant, so#

intGAD02minus <- glm (binaryGADbl ~ newsex + age + prev_psy + newFU0_Q35_4 + FU0_Q35_2 + FU0_Q35_3 + FU0_covid_status, data = data,

family = poisson (link = log))

intGAD02minus

coefs <- tidy(intGAD02minus, exponentiate = TRUE, conf.int = TRUE)

coefs

intGAD03 <- glm (binaryGADbl ~ newsex + age + prev_psy + newFU0_Q35_4 + FU0_Q35_2 + FU0_Q35_3 + FU0_covid_status + newFU0_Q35_4 * newsex, data = data,

family = poisson (link = log))

intGAD03

coefs <- tidy(intGAD03, exponentiate = TRUE, conf.int = TRUE)

coefs

menGAD03 <- glm (binaryGADbl ~ age + prev_psy + newFU0_Q35_4 + FU0_Q35_2 + FU0_Q35_3 + FU0_covid_status, data = dfmen,

family = poisson (link = log))

menGAD03

coefs<- tidy(menGAD03, exponentiate = TRUE, conf.int = TRUE)

coefs

womenGAD03 <- glm (binaryGADbl ~ age + prev_psy + newFU0_Q35_4 + FU0_Q35_2 + FU0_Q35_3 + FU0_covid_status, data = dfwomen,

family = poisson (link = log))

womenGAD03

coefs<- tidy(womenGAD03, exponentiate = TRUE, conf.int = TRUE)

coefs

intGAD61 <- glm (binaryGADhalf ~ newsex + age + prev_psy + newFU6_Q35_4 + FU6_Q35_2 + FU6_Q35_3 + FU6_covid_status + FU6_Q35_2 * newsex, data = data,

family = poisson (link = log))

intGAD61

coefs <- tidy(intGAD61, exponentiate = TRUE, conf.int = TRUE)

coefs #-> not significant, so#

intGAD61minus <- glm (binaryGADhalf ~ newsex + age + prev_psy + newFU6_Q35_4 + FU6_Q35_2 + FU6_Q35_3 + FU6_covid_status, data = data,

family = poisson (link = log))

intGAD61minus

coefs <- tidy(intGAD61minus, exponentiate = TRUE, conf.int = TRUE)

coefs

intGAD62 <- glm (binaryGADhalf ~ newsex + age + prev_psy + newFU6_Q35_4 + FU6_Q35_2 + FU6_Q35_3 + FU6_covid_status + FU6_Q35_3 * newsex, data = data,

family = poisson (link = log))

intGAD62

coefs <- tidy(intGAD62, exponentiate = TRUE, conf.int = TRUE)

coefs

menGAD62 <- glm (binaryGADhalf ~ age + prev_psy + newFU6_Q35_4 + FU6_Q35_2 + FU6_Q35_3 + FU6_covid_status, data = dfmen,

family = poisson (link = log))

menGAD62

coefs<- tidy(menGAD62, exponentiate = TRUE, conf.int = TRUE)

coefs

womenGAD62 <- glm (binaryGADhalf ~ age + prev_psy + newFU6_Q35_4 + FU6_Q35_2 + FU6_Q35_3 + FU6_covid_status, data = dfwomen,

family = poisson (link = log))

womenGAD62

coefs<- tidy(womenGAD62, exponentiate = TRUE, conf.int = TRUE)

coefs

intGAD63 <- glm (binaryGADhalf ~ newsex + age + prev_psy + newFU6_Q35_4 + FU6_Q35_2 + FU6_Q35_3 + FU6_covid_status + newFU6_Q35_4 * newsex, data = data,

family = poisson (link = log))

intGAD63

coefs <- tidy(intGAD63, exponentiate = TRUE, conf.int = TRUE)

coefs#-> not significant, so#

intGAD63minus <- glm (binaryGADhalf ~ newsex + age + prev_psy + newFU6_Q35_4 + FU6_Q35_2 + FU6_Q35_3 + FU6_covid_status, data = data,

family = poisson (link = log))

intGAD63minus

coefs <- tidy(intGAD63minus, exponentiate = TRUE, conf.int = TRUE)

coefs

intGAD121 <- glm (binaryGADone ~ newsex + age + prev_psy + newFU12_Q35_4 + FU12_Q35_2 + FU12_Q35_3 + FU12_covid_status + FU12_Q35_2 * newsex, data = data,

family = poisson (link = log))

intGAD121

coefs <- tidy(intGAD121, exponentiate = TRUE, conf.int = TRUE)

coefs#-> not significant, so#

intGAD121minus <- glm (binaryGADone ~ newsex + age + prev_psy + newFU12_Q35_4 + FU12_Q35_2 + FU12_Q35_3 + FU12_covid_status, data = data,

family = poisson (link = log))

intGAD121minus

coefs <- tidy(intGAD121minus, exponentiate = TRUE, conf.int = TRUE)

coefs

intGAD122 <- glm (binaryGADone ~ newsex + age + prev_psy + newFU12_Q35_4 + FU12_Q35_2 + FU12_Q35_3 + FU12_covid_status + FU12_Q35_3 * newsex, data = data,

family = poisson (link = log))

intGAD122

coefs <- tidy(intGAD122, exponentiate = TRUE, conf.int = TRUE)

coefs#-> not significant, so#

intGAD122minus <- glm (binaryGADone ~ newsex + age + prev_psy + newFU12_Q35_4 + FU12_Q35_2 + FU12_Q35_3 + FU12_covid_status, data = data,

family = poisson (link = log))

intGAD122minus

coefs <- tidy(intGAD122minus, exponentiate = TRUE, conf.int = TRUE)

coefs

intGAD123 <- glm (binaryGADone ~ newsex + age + prev_psy + newFU12_Q35_4 + FU12_Q35_2 + FU12_Q35_3 + FU12_covid_status + newFU12_Q35_4 * newsex, data = data,

family = poisson (link = log))

intGAD123

coefs <- tidy(intGAD123, exponentiate = TRUE, conf.int = TRUE)

coefs#-> not significant, so#

intGAD123minus <- glm (binaryGADone ~ newsex + age + prev_psy + newFU12_Q35_4 + FU12_Q35_2 + FU12_Q35_3 + FU12_covid_status, data = data,

family = poisson (link = log))

intGAD123minus

coefs <- tidy(intGAD123minus, exponentiate = TRUE, conf.int = TRUE)

coefs

intptsd01 <- glm (binaryptsd0 ~ newsex + age + prev_psy + newFU0_Q35_4 + FU0_Q35_2 + FU0_Q35_3 + FU0_covid_status + FU0_Q35_2 * newsex, data = data,

family = poisson (link = log))

intptsd01

coefs <- tidy(intptsd01, exponentiate = TRUE, conf.int = TRUE)

coefs

menptsd01 <- glm (binaryptsd0 ~ age + prev_psy + newFU0_Q35_4 + FU0_Q35_2 + FU0_Q35_3 + FU0_covid_status, data = dfmen,

family = poisson (link = log))

menptsd01

coefs<- tidy(menptsd01, exponentiate = TRUE, conf.int = TRUE)

coefs

womenptsd01 <- glm (binaryptsd0 ~ age + prev_psy + newFU0_Q35_4 + FU0_Q35_2 + FU0_Q35_3 + FU0_covid_status, data = dfwomen,

family = poisson (link = log))

womenptsd01

coefs<- tidy(womenptsd01, exponentiate = TRUE, conf.int = TRUE)

coefs

intptsd02 <- glm (binaryptsd0 ~ newsex + age + prev_psy + newFU0_Q35_4 + FU0_Q35_2 + FU0_Q35_3 + FU0_covid_status + FU0_Q35_3 * newsex, data = data,

family = poisson (link = log))

intptsd02

coefs <- tidy(intptsd02, exponentiate = TRUE, conf.int = TRUE)

coefs#-> not significant, so#

intptsd02minus <- glm (binaryptsd0 ~ newsex + age + prev_psy + newFU0_Q35_4 + FU0_Q35_2 + FU0_Q35_3 + FU0_covid_status, data = data,

family = poisson (link = log))

intptsd02minus

coefs <- tidy(intptsd02minus, exponentiate = TRUE, conf.int = TRUE)

coefs

intptsd03 <- glm (binaryptsd0 ~ newsex + age + prev_psy + newFU0_Q35_4 + FU0_Q35_2 + FU0_Q35_3 + FU0_covid_status + newFU0_Q35_4 * newsex, data = data,

family = poisson (link = log))

intptsd03

coefs <- tidy(intptsd03, exponentiate = TRUE, conf.int = TRUE)

coefs#-> not significant, so#

intptsd03minus <- glm (binaryptsd0 ~ newsex + age + prev_psy + newFU0_Q35_4 + FU0_Q35_2 + FU0_Q35_3 + FU0_covid_status, data = data,

family = poisson (link = log))

intptsd03minus

coefs <- tidy(intptsd03minus, exponentiate = TRUE, conf.int = TRUE)

coefs

intptsd61 <- glm (binaryptsd6 ~ newsex + age + prev_psy + newFU6_Q35_4 + FU6_Q35_2 + FU6_Q35_3 + FU6_covid_status + FU6_Q35_2 * newsex, data = data,

family = poisson (link = log))

intptsd61

coefs<- tidy(intptsd61, exponentiate = TRUE, conf.int = TRUE)

coefs#-> not significant, so#

intptsd61minus <- glm (binaryptsd6 ~ newsex + age + prev_psy + newFU6_Q35_4 + FU6_Q35_2 + FU6_Q35_3 + FU6_covid_status, data = data,

family = poisson (link = log))

intptsd61minus

coefs <- tidy(intptsd61minus, exponentiate = TRUE, conf.int = TRUE)

coefs

intptsd62 <- glm (binaryptsd6 ~ newsex + age + prev_psy + newFU6_Q35_4 + FU6_Q35_2 + FU6_Q35_3 + FU6_covid_status + FU6_Q35_3 * newsex, data = data,

family = poisson (link = log))

intptsd62

coefs<- tidy(intptsd62, exponentiate = TRUE, conf.int = TRUE)

coefs

menptsd62 <- glm (binaryptsd6 ~ age + prev_psy + newFU6_Q35_4 + FU6_Q35_2 + FU6_Q35_3 + FU6_covid_status, data = dfmen,

family = poisson (link = log))

menptsd62

coefs<- tidy(menptsd62, exponentiate = TRUE, conf.int = TRUE)

coefs

womenptsd62 <- glm (binaryptsd6 ~ age + prev_psy + newFU6_Q35_4 + FU6_Q35_2 + FU6_Q35_3 + FU6_covid_status, data = dfwomen,

family = poisson (link = log))

womenptsd62

coefs<- tidy(womenptsd62, exponentiate = TRUE, conf.int = TRUE)

coefs

intptsd63 <- glm (binaryptsd6 ~ newsex + age + prev_psy + newFU6_Q35_4 + FU6_Q35_2 + FU6_Q35_3 + FU6_covid_status + newFU6_Q35_4 * newsex, data = data,

family = poisson (link = log))

intptsd63

coefs<- tidy(intptsd63, exponentiate = TRUE, conf.int = TRUE)

coefs#-> not significant, so#

intptsd63minus <- glm (binaryptsd6 ~ newsex + age + prev_psy + newFU6_Q35_4 + FU6_Q35_2 + FU6_Q35_3 + FU6_covid_status, data = data,

family = poisson (link = log))

intptsd63minus

coefs <- tidy(intptsd63minus, exponentiate = TRUE, conf.int = TRUE)

coefs

intptsd121 <- glm (binaryptsd12 ~ newsex + age + prev_psy + newFU12_Q35_4 + FU12_Q35_2 + FU12_Q35_3 + FU12_covid_status + FU12_Q35_2 * newsex, data = data,

family = poisson (link = log))

intptsd121

coefs<- tidy(intptsd121, exponentiate = TRUE, conf.int = TRUE)

coefs#-> not significant, so#

intptsd121minus <- glm (binaryptsd12 ~ newsex + age + prev_psy + newFU12_Q35_4 + FU12_Q35_2 + FU12_Q35_3 + FU12_covid_status, data = data,

family = poisson (link = log))

intptsd121minus

coefs <- tidy(intptsd121minus, exponentiate = TRUE, conf.int = TRUE)

coefs

intptsd122 <- glm (binaryptsd12 ~ newsex + age + prev_psy + newFU12_Q35_4 + FU12_Q35_2 + FU12_Q35_3 + FU12_covid_status + FU12_Q35_3 * newsex, data = data,

family = poisson (link = log))

intptsd122

coefs<- tidy(intptsd122, exponentiate = TRUE, conf.int = TRUE)

coefs#-> not significant, so#

intptsd122minus <- glm (binaryptsd12 ~ newsex + age + prev_psy + newFU12_Q35_4 + FU12_Q35_2 + FU12_Q35_3 + FU12_covid_status, data = data,

family = poisson (link = log))

intptsd122minus

coefs <- tidy(intptsd122minus, exponentiate = TRUE, conf.int = TRUE)

coefs

intptsd123 <- glm (binaryptsd12 ~ newsex + age + prev_psy + newFU12_Q35_4 + FU12_Q35_2 + FU12_Q35_3 + FU12_covid_status + newFU12_Q35_4 * newsex, data = data,

family = poisson (link = log))

intptsd123

coefs<- tidy(intptsd123, exponentiate = TRUE, conf.int = TRUE)

coefs#-> not significant, so#

intptsd123minus <- glm (binaryptsd12 ~ newsex + age + prev_psy + newFU12_Q35_4 + FU12_Q35_2 + FU12_Q35_3 + FU12_covid_status, data = data,

family = poisson (link = log))

intptsd123minus

coefs <- tidy(intptsd123minus, exponentiate = TRUE, conf.int = TRUE)

coefs

#stratified by gender#####

menPHQ0 <- glm (binaryPHQbl ~ age + prev_psy + newFU0_Q35_4 + FU0_Q35_2 + FU0_Q35_3 + FU0_covid_status + FU0_Q35_3 * FU0_Q35_2, data = dfmen,

family = poisson (link = log))

menPHQ0

coefs<- tidy(menPHQ0, exponentiate = TRUE, conf.int = TRUE)

coefs

menPHQ6 <- glm (binaryPHQhalf ~ age + prev_psy + newFU6_Q35_4 + FU6_Q35_2 + FU6_Q35_3 + FU6_covid_status + FU6_Q35_3 * FU6_Q35_2, data = dfmen,

family = poisson (link = log))

menPHQ6

coefs<- tidy(menPHQ6, exponentiate = TRUE, conf.int = TRUE)

coefs

menPHQ12 <- glm (binaryPHQone ~ age + prev_psy + newFU12_Q35_4 + FU12_Q35_2 + FU12_Q35_3 + FU12_covid_status + FU12_Q35_3 * FU12_Q35_2, data = dfmen,

family = poisson (link = log))

menPHQ12

coefs<- tidy(menPHQ12, exponentiate = TRUE, conf.int = TRUE)

coefs

menGAD0 <- glm (binaryGADbl ~ age + prev_psy + newFU0_Q35_4 + FU0_Q35_2 + FU0_Q35_3 + FU0_covid_status + FU0_Q35_3 * FU0_Q35_2, data = dfmen,

family = poisson (link = log))

menGAD0

coefs<- tidy(menGAD0, exponentiate = TRUE, conf.int = TRUE)

coefs

menGAD6 <- glm (binaryGADhalf ~ age + prev_psy + newFU6_Q35_4 + FU6_Q35_2 + FU6_Q35_3 + FU6_covid_status + FU6_Q35_3 * FU6_Q35_2, data = dfmen,

family = poisson (link = log))

menGAD6

coefs<- tidy(menGAD6, exponentiate = TRUE, conf.int = TRUE)

coefs

menGAD12 <- glm (binaryGADone ~ age + prev_psy + newFU12_Q35_4 + FU12_Q35_2 + FU12_Q35_3 + FU12_covid_status + FU12_Q35_3 * FU12_Q35_2, data = dfmen,

family = poisson (link = log))

menGAD12

coefs<- tidy(menGAD12, exponentiate = TRUE, conf.int = TRUE)

coefs

menptsd0 <- glm (binaryptsd0 ~ age + prev_psy + newFU0_Q35_4 + FU0_Q35_2 + FU0_Q35_3 + FU0_covid_status + FU0_Q35_3 * FU0_Q35_2, data = dfmen,

family = poisson (link = log))

menptsd0

coefs<- tidy(menptsd0, exponentiate = TRUE, conf.int = TRUE)

coefs

menptsd6 <- glm (binaryptsd6 ~ age + prev_psy + newFU6_Q35_4 + FU6_Q35_2 + FU6_Q35_3 + FU6_covid_status + FU6_Q35_3 * FU6_Q35_2, data = dfmen,

family = poisson (link = log))

menptsd6

coefs<- tidy(menptsd6, exponentiate = TRUE, conf.int = TRUE)

coefs

menptsd12 <- glm (binaryptsd12 ~ age + prev_psy + newFU12_Q35_4 + FU12_Q35_2 + FU12_Q35_3 + FU12_covid_status + FU12_Q35_3 * FU12_Q35_2, data = dfmen,

family = poisson (link = log))

menptsd12

coefs<- tidy(menptsd12, exponentiate = TRUE, conf.int = TRUE)

coefs

womenPHQ0 <- glm (binaryPHQbl ~ age + prev_psy + newFU0_Q35_4 + FU0_Q35_2 + FU0_Q35_3 + FU0_covid_status + FU0_Q35_3 * FU0_Q35_2, data = dfwomen,

family = poisson (link = log)).

womenPHQ0

coefs<- tidy(womenPHQ0, exponentiate = TRUE, conf.int = TRUE)

coefs

womenPHQ6 <- glm (binaryPHQhalf ~ age + prev_psy + newFU6_Q35_4 + FU6_Q35_2 + FU6_Q35_3 + FU6_covid_status + FU6_Q35_3 * FU6_Q35_2, data = dfwomen,

family = poisson (link = log))

womenPHQ6

coefs<- tidy(womenPHQ6, exponentiate = TRUE, conf.int = TRUE)

coefs

womenPHQ12 <- glm (binaryPHQone ~ age + prev_psy + newFU12_Q35_4 + FU12_Q35_2 + FU12_Q35_3 + FU12_covid_status + FU12_Q35_3 * FU12_Q35_2, data = dfwomen,

family = poisson (link = log))

womenPHQ12

coefs<- tidy(womenPHQ12, exponentiate = TRUE, conf.int = TRUE)

coefs

womenGAD0 <- glm (binaryGADbl ~ age + prev_psy + newFU0_Q35_4 + FU0_Q35_2 + FU0_Q35_3 + FU0_covid_status + FU0_Q35_3 * FU0_Q35_2, data = dfwomen,

family = poisson (link = log))

womenGAD0

coefs<- tidy(womenGAD0, exponentiate = TRUE, conf.int = TRUE)

coefs

womenGAD6 <- glm (binaryGADhalf ~ age + prev_psy + newFU6_Q35_4 + FU6_Q35_2 + FU6_Q35_3 + FU6_covid_status + FU6_Q35_3 * FU6_Q35_2, data = dfwomen,

family = poisson (link = log))

womenGAD6

coefs<- tidy(womenGAD6, exponentiate = TRUE, conf.int = TRUE)

coefs

womenGAD12 <- glm (binaryGADone ~ age + prev_psy + newFU12_Q35_4 + FU12_Q35_2 + FU12_Q35_3 + FU12_covid_status + FU12_Q35_3 * FU12_Q35_2, data = dfwomen,

family = poisson (link = log))

womenGAD12

coefs<- tidy(womenGAD12, exponentiate = TRUE, conf.int = TRUE)

coefs

womenptsd0 <- glm (binaryptsd0 ~ age + prev_psy + newFU0_Q35_4 + FU0_Q35_2 + FU0_Q35_3 + FU0_covid_status + FU0_Q35_3 * FU0_Q35_2, data = dfwomen,

family = poisson (link = log))

womenptsd0

coefs<- tidy(womenptsd0, exponentiate = TRUE, conf.int = TRUE)

coefs

womenptsd6 <- glm (binaryptsd6 ~ age + prev_psy + newFU6_Q35_4 + FU6_Q35_2 + FU6_Q35_3 + FU6_covid_status + FU6_Q35_3 * FU6_Q35_2, data = dfwomen,

family = poisson (link = log))

womenptsd6

coefs<- tidy(womenptsd6, exponentiate = TRUE, conf.int = TRUE)

coefs

womenptsd12 <- glm (binaryptsd12 ~ age + prev_psy + newFU12_Q35_4 + FU12_Q35_2 + FU12_Q35_3 + FU12_covid_status + FU12_Q35_3 * FU12_Q35_2, data = dfwomen,

family = poisson (link = log))

womenptsd12

coefs<- tidy(womenptsd12, exponentiate = TRUE, conf.int = TRUE)

coefs

- *Worry about delayed care was measured on a scale with multiple options at baseline. This was later corrected with means been calculated.*
